# Multi-omic Characterization of the Oral Ecosystem and Assessment of Dental Health and Dental Hygiene Metadata in the Participants from a Large Randomized Double-Blinded Clinical Study

**DOI:** 10.64898/2026.09.24.26363963

**Authors:** Hannah Moglowsky, Sofronia M. Ringold, Rodney Jones, Natalie Wall, Alexander Gudjonsson, Thorsten Gravert, Janne Marie Moll, Anne O’Donnell, Katie Lee, Emily A. Stein

## Abstract

Increasing evidence highlights the important role the oral ecosystem plays in systemic health. Despite the fact that oral diseases affect nearly 3.5 billion people globally, our understanding of the precise drivers of these diseases remains limited. To bridge this gap and provide broader understanding of oral public health, a clinical study involving 595 participants 20-78 years of age was conducted over 8 weeks. Characterization of the oral ecosystem using metabolomics, metagenomics, and targeted immune markers were analyzed in conjunction with demographic, lifestyle, and medical history metadata. This comprehensive approach allowed for the identification of specific links between microbial and metabolic signatures and overall wellness, providing a contemporary baseline to enable the development of personalized prevention and treatment strategies to help safeguard oral and systemic public health.

## Introduction

The global burden of oral disease is approaching 3.5 billion people [1]. Despite the recognition of oral health as a pillar of systemic well-being and significant advances in dental technologies, health disparities remain, particularly among socioeconomically disadvantaged and racially marginalized populations who face complex barriers to care [2]. For example, national data reveals that the prevalence of severe periodontitis is significantly higher among Black and Hispanic adults compared to Caucasian adults [3,4]. In 2021, 20.1% of Minnesotans reported not seeking health care, including dental, due to cost or lack of insurance [5,6]. These populations are particularly at risk of poor dental health, leading to a “silent epidemic” of permanent tooth loss and untreated decay, which can impact learning, earning potential, lifespan and quality of life [7–9].

Conceptualization of human health has undergone a paradigm shift towards a holistic understanding of the body as a complex “holobiont,” a unified biological system comprising human cells and a vast, diverse community of microorganisms [10]. Previous research has shown that the mouth harbors the second-most diverse microbial population in the human body with over 700 bacterial species [10,11]. Health is theorized to result in an overall balanced microbiome, the opposite of which is driven by imbalance, with growing evidence suggesting a key driver may be microbial dysregulation (dysbiosis) [10,11]. Within this framework, tooth remineralization, digestion, mucosal integrity, and immune response can be impacted by metabolic, microbial, lifestyle, and/or environmental changes [1,12,13].

Traditional views of oral diseases, dental caries, and periodontal disease as localized conditions are being replaced by the recognition that these can be chronic, multifactorial diseases inextricably linked to microbial dysbiosis, systemic inflammation, metabolic dysfunction, lifestyle, and other factors [11,14–17].

Application of high-resolution multi-omics platforms – including metagenomics, metabolomics and host inflammatory profiling – to characterize oral inflammatory mediators, metabolic signatures and microbial taxa, and their functional pathways will enhance understanding of the interactions between oral hygiene behaviors and the oral ecosystem. This integrative approach enables identification of predictive biomarkers and risk factors that drive the pathogenesis of dental diseases, thereby informing precision public oral health strategies.

This study was designed to address these unmet needs by evaluating a representative population from the Minneapolis-St. Paul metro area through a comprehensive, 8-week randomized controlled trial. The first objective was to characterize 595 volunteers’ oral metabolomes, oral metagenomes, salivary immune markers, medical histories, oral hygiene habits, dietary patterns, exercise frequency, and emotional stressors to gain a better understanding of the relationship between multiple variables that relate to public health. The second objective was to identify variables of inter-dependence that relate to specific features found in the metadata, metagenomic, metabolomic, and/or immune marker data to provide an improved understanding of the relationship between lifestyle behaviors and sophisticated biomarkers that relate to dental health and/or diseases.

The overarching hypothesis driving this study is that distinct oral multi-omic signatures captured through metabolomics, metagenomics and salivary immune profiling are significantly associated with self-reported behaviors and medical history. These signatures may reveal latent risk profiles for chronic disease that are not detectable through traditional clinical assessments.

The overall aim of this work was to establish a more thorough understanding of the variances within population health to identify significant behaviors and physical features that associate with health in an urban community.

## Materials and Methods

### Study Design and Ethics Approval

This study includes an analysis of the baseline data collected as part of an 8-week, three-armed, randomized, controlled, double-blinded intervention trial (ClinicalTrials.gov registration number NCT07451782), conducted in compliance with the International Council for Harmonization Guideline on Good Clinical Practices (ICH-GCP) and adhering to the principles outlined in the Declaration of Helsinki. Participants were recruited between April 2024 and July 2024. The study procedures spanned 60 days of intervention and clinical research activities were carried out in the Minneapolis-Saint Paul metro area, Minnesota, USA. The study protocol was reviewed and approved by Advarra IRB (Approval Number: Pro 00078044), and all participants provided written informed consent prior to enrollment.

### Study Population and Selection Criteria

A total of 595 participants (targeted a maximum of 600) were enrolled from the Minneapolis-Saint Paul metro area by Comprehensive Research Group (Minneapolis, MN). The study focused on individuals representing a broad spectrum of demographics, including age (18-80 years) and diverse economic and social statuses. Key inclusion criteria required participants to be non-smokers for a minimum of two years and to have had no oral antibiotic used within 30 days prior to enrollment. Exclusion criteria included smoking, pregnancy, breastfeeding, or recent antibiotics due to their established impact on the metabolome and metagenome [18–22]. Participants also could not have recently participation in another oral clinical trial. Sample size was determined to reflect a clinically relevant effect size to capture a diverse dataset to power statistical analyses.

### Self-Reported Questionnaire Data Acquisition

The study consisted of three scheduled visits at Baseline (Visit 1), Week 4 (Visit 2), and Week 8 (Visit 3). At each of the three visits, participants completed self-reported questionnaires on a study-provided laptop covering medical, dental, diet, exercise, and stress (Perceived Stress Questionnaire-30) [23,24]. The PSQ-30 is a validated questionnaire to assess subjective stress [23,24]. To be more specific, the context “at home”/ “at your job” was added to items 2 and 6 of the PSQ-30 for a total of 32 questions (PSQ-32). A total score, index score, and dimension scores (i.e., lack of joy, harassment, irritability, overload, fatigue, tension, and worries) were calculated. Questionnaire responses were linked to participant data only by a unique, non-identifiable code.

### Study Procedures and Sample Collection

Participants were required to refrain from eating, drinking, or using any oral hygiene products for one hour prior to sample collection. At each visit, a qualified technician collected five oral samples per participant, resulting in 1,684 total samples analyzed. The sampling used 6” sterile flocked swabs (Puritan Medical Products). The swabs were collected from the tooth-gingival interface across four distinct quadrants of the mouth, plus a final “resting control swab” to capture the sampling environment at that clinical site, as a control for sample collection on that day. In total, seven batches of swabs were collected over the period of 60 days.

For each biological sample, two swabs were collected and combined into a single sterile 15-ml conical tube (Corning, 430790). Immediately after collection, tubes were flash frozen and stored at -80°C until shipment on dry ice. Sterile flocked swabs designated for shotgun metagenomic analysis were processed at Cmbio (Germantown, Maryland, USA). Swabs designated for Metabolomic analysis were collected into special tubes (Salimetrics, 5001.05) and processed at Cmbio (København, Denmark).

### Metabolomic Data Processing and Analysis

Prior to study initiation, fresh and frozen oral swab samples were tested to confirm optimal sample recovery for metabolome evaluation (data not shown). The extraction protocol work-up was performed with pooled quality control samples to maximize batch-to-batch comparability to aid in normalization across all 7 batches. After feature extraction, grouping and integration, signal measurements were matched against references, metabolite annotation confidence was classified in four levels with level 1 - highest confidence to level 3 - lower confidence (Suppl. Fig. 1). Signals were then filtered by calculating quality control metrics and predefined thresholds i.e. signal-to-noise (above 3), coefficient of variance representing precision (below 20%), dilution-to-signal correlation (above 0.5), and mean QC signal-to-mean sample signal (between 0.5 and 2.0) were established (Suppl. Fig. 1).

Metabolomic analysis included both semi-polar untargeted profiling and targeted quantification performed by Cmbio (Vedbæk, Denmark). Saliva was first removed from swabs by centrifugation (3,000 x g for 15 minutes at 4°C). Swabs were cut and transferred to Eppendorf tubes, and an extraction protocol was implemented using an extraction solvent (150 μL, 10 mM ammonium formate in water/methanol 9:1, v/v, containing 0.1% formic acid), supplemented with stable isotope-labeled internal standards. Samples were homogenized using a VWR® Star-Beater (30 Hz for 2 minutes). The homogenized samples were filtered through a Spin-X® centrifuge tube filter (22 μM, Corning® Costar®) by centrifugation (15,000 x g for 5 minutes at 4°C) prior to instrumental analysis.

Semi-polar metabolite profiling was performed using a Vanquish Ultra-High-Performance Liquid Chromatography (UHPLC) system coupled to a Thermo Scientific Orbitrap Exploris 240 mass spectrometer (MS) with a heated electrospray ionization (ESI) source. Chromatographic separation utilized an ACQUITY UPLC HSS T3 column (1.8 μM, 2.1 mm x 150 mm; Waters), following an adapted version of the method described by Doneanu et al. (2011) [25]. The mobile phase consisted of Eluent A (10 mM ammonium formate in water with 0.1% formic acid) and Eluent B (10 mM ammonium formate in methanol with 0.1% formic acid). Data were in full MS/dd-MS2 mode with a scan range of 50-750 m/z. Full MS resolution was set to 60,000, while dd-MS2 resolution was set to 30,000 with a Top N precursor selection of 5. An isolation window of 0.4 m/z was utilized alongside stepped collision energy (NCE) settings of 20, 40, and 60.

Raw data were processed using Compound Discoverer 3.3 (Thermo Scientific), Skyline 24.1 (MacCoss Lab) and MATLAB (MathWorks). L-Citrulline (HMDB0000904), Trimethylamine (HMDB0000906) and Trimethylamine N-oxide (HMDB0000925) were extracted, targeted, and quantified against external calibration curves of authentic reference standards (40.00 μM to 0.004 μM). Compound annotation confidence adhered to community standards: Level 1 (accurate mass, MS/MS spectra, and known retention time); Level 2a (accurate mass and retention time); Level 2b (accurate mass and MS/MS spectra matching public databases); and Level 3 (accurate mass and elemental composition). Annotations were supported by in-house libraries and public databases, including HMDB v5.0 and the Saliva Metabolome Database [26].

Data preprocessing and normalization procedures were tailored to each omics layer. For metabolomics, 2,245 injections were processed across seven analytical batches, including 1,682 study samples, 215 quality control (QC) samples, and 46 blanks. Two types of QC samples were utilized “QC_Global” (aliquots from the entire sample set) and “QC_Batch” (aliquots from each specific batch). QC samples were injected at the beginning, end, and every 10 samples throughout the sequence to monitor drift and normalize batch effects. Three QC_Global dilution series were prepared to evaluate detector response linearity. Missing values in untargeted analysis were filled using Compound Discoverer’s Gap Fill Node, while targeted metabolite peak areas were imputed using random numbers below instrument noise level. Analytical batch variation was corrected using Systematic Error Removal using Random Forest (SERRF), with separate normalization for study/QC samples versus blanks. Biological dilution variation was corrected using Probabilistic Quotient Normalization (PQN) [27].

For each data type, the undetected values were discarded from continuous analyses but included in binomial detection analyses. Metabolites whose peak areas measured above the compound specific limit of detection were considered detected and their PQN peak area was power transformed for continuous analyses. Immunological biomarkers whose concentrations measured above the marker specific limit of detection were considered detected and their concentrations log_2_-transformed for continuous analyses.

### Metagenomic Data Processing and Analysis

Prior to the clinical study, microbial DNA load per oral swab was examined. Two swab samples were collected per subject to reduce sampling variability and increase recovery during extraction (data not shown). 99.8% of samples met acceptance criteria (6 Gbp ∼20M read pairs at an average of 35.4M read pairs per sample, minimum of 12.4M read pairs). As expected, an average of 88.1% of all reads were host DNA; however, a significant proportion of reads were high-quality non-host. 96.8% of samples met the quality criteria having a 95% probability at which the relative abundance of an average species would be detected below the threshold of 0.1% limit of detection and were used in downstream analyses (Suppl Fig. 2A-D). Five baseline samples did not meet the sequencing threshold and were excluded from further metagenomic analysis.

DNA was extracted from oral swabs using the QIAGEN DNeasy PowerSoil Pro Kit. DNA libraries were prepared using the Watchmaker DNA Library Prep Kit with IDT xGen UDI Primers and Stubby Adapters, followed by 7 cycles of PCR. Shotgun metagenomic sequencing was performed on an Illumina NovaSeq X platform, yielding paired end reads of 2 x 150 bp, with an average of 35.4 million read pairs per sample. Raw reads were trimmed to remove adapters and bases with a Phred score below 30 using AdapterRemoval v.2.3.1. Human host DNA contamination was removed by mapping reads against the human reference genome (GRCh38.p14) with Bowtie2 v.2.4.2. High-quality non-host (HQNH) reads were retained for downstream analysis. Samples with fewer than 5,000 reads mapping to signature genes were excluded from all analyses. Species-level relative abundances were estimated using the Cmbio Human Microbiome Profiler (CHAMP, Pita et al. 2024) against the Cmbio Human Microbiome Reference HMR05 gene catalog. Taxonomic annotation was based on the Genome Taxonomy Database (GTDB) release 214. Functional potential was profiled using Kyoto Encyclopedia of Genes and Genomes (KEGG) modules (v. 78.2). Rarefied relative abundance data was obtained by random sampling 10,000 signature reads per sample.

Metagenomic species with a rarefied relative abundance above zero were considered detected and for continuous analyses the non-zero non-rarefied relative abundance was log_2_-transformed. All values for continuous analyses were winsorized at the 0.03 and 0.97 quantiles. Taxa and metabolites present in at least 10% of subjects were considered in the analysis.

### Multi-variate and Statistical Analyses

All multi-omic and immune marker analyses were conducted on baseline samples from 595 participants. Behavioral data analysis was conducted on the self-reported questionnaires from each visit. The primary analysis characterized the oral metabolome, microbiome, and immunological biomarkers.

Dimensionality reduction (PCA, PCoA) and diversity assessment (Richness and Weighted UniFrac) were employed to visualize clustering and diversity. Oral community type (OCT) clustering was performed using Dirichlet multinomial mixture modeling [28] based on species signature gene counts (N=590). To analyze the dispersion of the OCTs in Weighted UniFrac beta diversity space, distances from individual samples to their respective OCTs centroid were compared using a permutational test of multivariate homogeneity of group dispersions. Associations between microbiome taxon abundance, metabolite intensity, immune biomarkers, and questionnaire features (111 items) were tested using linear regression with LinDA compositional bias correction for microbiome data [29]. Associations between microbiome taxon abundance, metabolite intensity, immune biomarkers, and questionnaire features (111 items) and OCT assignments were assessed with multinomial regression models weighted by the Dirichlet parameter. To perform association analyses with immunological biomarkers, the ng/mL levels of protein determined by ELISA were log-transformed to enable multinomial regression. All P-values were adjusted using the Benjamini-Hochberg method (FDR<0.1). Statistical analyses were performed using R [30].

### Outcomes

The primary outcome was comprehensive characterization of the oral metabolome, oral microbiome, and the salivary immunological biomarkers interleukin-6 (IL-6) and matrix metalloproteinases 8 and 13 (MMP-8, MMP-13, respectively) in a population representative of the Minneapolis-St. Paul metro area. Metabolome analysis included untargeted and targeted metabolite detections based on UHPLC coupled to high resolution tandem Mass Spectrometry. Microbiome profiling encompassed taxonomic and functional annotation, alpha diversity, and beta diversity.

Secondary outcomes assessed the cross-sectional differences in the integrated multi-omic profiles in participants with self-reported differences in dental hygiene practices, dietary habits and age.

## Results

### Study Participants

The final sample included 595 participants from the Minneapolis-St. Paul metro area, 87% of which were female, aged 20-78 (mean=54.45 ± 13.86; (Table 1). 68.7% identified as White or Caucasian, 16.5% as Black or African American, 5.9% as Hispanic or Latino, and 8.9% as Other/Multiracial. 595 participants completed the baseline visit, 590 completed week 4, and 531 completed week 8.

**Table 1.** Study Population Demographics.

| Characteristic | Total Population (N=595) |
| --- | --- |
| **Age (Years)\*** |  |
| Mean [SD] | 54.45 [13.86] |
| Median [Min, Max] | 57 [20, 78] |

| **Sex** |  |
| --- | --- |
| Female | 522 (87.7%) |
| Male | 73 (12.3%) |
| **Race/Ethnicity** |  |
| White or Caucasian | 409 (68.7%) |
| Black or African American | 98 (16.5%) |
| Hispanic or Latino | 35 (5.9%) |
| Other/Multiracial | 53 (8.9%) |

\* One participant (Subject 1323) year of birth was 2024, a data entry error; this subject was excluded from age calculations.

### Medical History and Medication Usage

Medical diagnoses with a prevalence at or above 5% of the study population are listed in Table 2, with the full list in Supplementary Table S1. Anxiety was the most prevalent condition reported (19.3%), followed by headaches (18.5%), and then arthritis (17.8%). 60.2% of participants reported taking medications (Table 2), with Supplements & Vitamins as the most common (37%), followed by Statins and antihyperlipidemics (13.8%) and thyroid hormones (10.8%; Table 3). The full list of medications by category is listed in Supplemental Table S2. The mean PSQ-32 raw score was 55.17 ± 15.85 and the mean index score was 0.25 ±0.17.

**Table 2.** Medical History, Medication Usage, and PSQ-32 Scores.

| **Medical History [$\geq 5\%$ Prevalence] (N=595)** | **Yes N (%)** |
| --- | --- |
| Anxiety | 115 (19.3%) |
| Headaches | 110 (18.5%) |
| Arthritis | 106 (17.8%) |
| High Blood Pressure | 103 (17.3%) |
| Depression | 97 (16.3%) |
| High Cholesterol | 94 (15.8%) |
| Impaired Vision | 93 (15.6%) |
| Acid Reflux/GERD | 70 (11.8%) |
| Thyroid Problem | 59 (9.9%) |
| Sleep Apnea | 55 (9.2%) |
| Asthma | 46 (7.7%) |
| Sinus Inflammation | 41 (6.9%) |
| Other Skin Condition | 41 (6.9%) |
| Hearing Loss | 40 (6.7%) |
| Cancer/Tumor | 34 (5.7%) |
| Tinnitus | 34 (5.7%) |
| Irritable Bowel Syndrome | 30 (5.0%) |
| **PSQ-32 (N=531)** | **Mean (SD)** |
| PSQ_Total | 55.71 (15.85) |
| PSQ_Index | 0.25 (0.17) |
| PSQ_Harassment | 1.50 (0.48) |
| PSQ_Overload | 2.01 (0.72) |
| PSQ_Irritability | 1.72 (0.59) |
| PSQ_LackOfJoy | 1.72 (0.53) |
| PSQ_Fatigue | 2.01 (0.63) |
| PSQ_Worries | 1.64 (0.60) |
| PSQ_Tension | 1.74 (0.58) |
| **Medication Usage (N=595)** | **Yes N (%)** |
| Takes Medications | 358 (60.2%) |

**Table 3:**
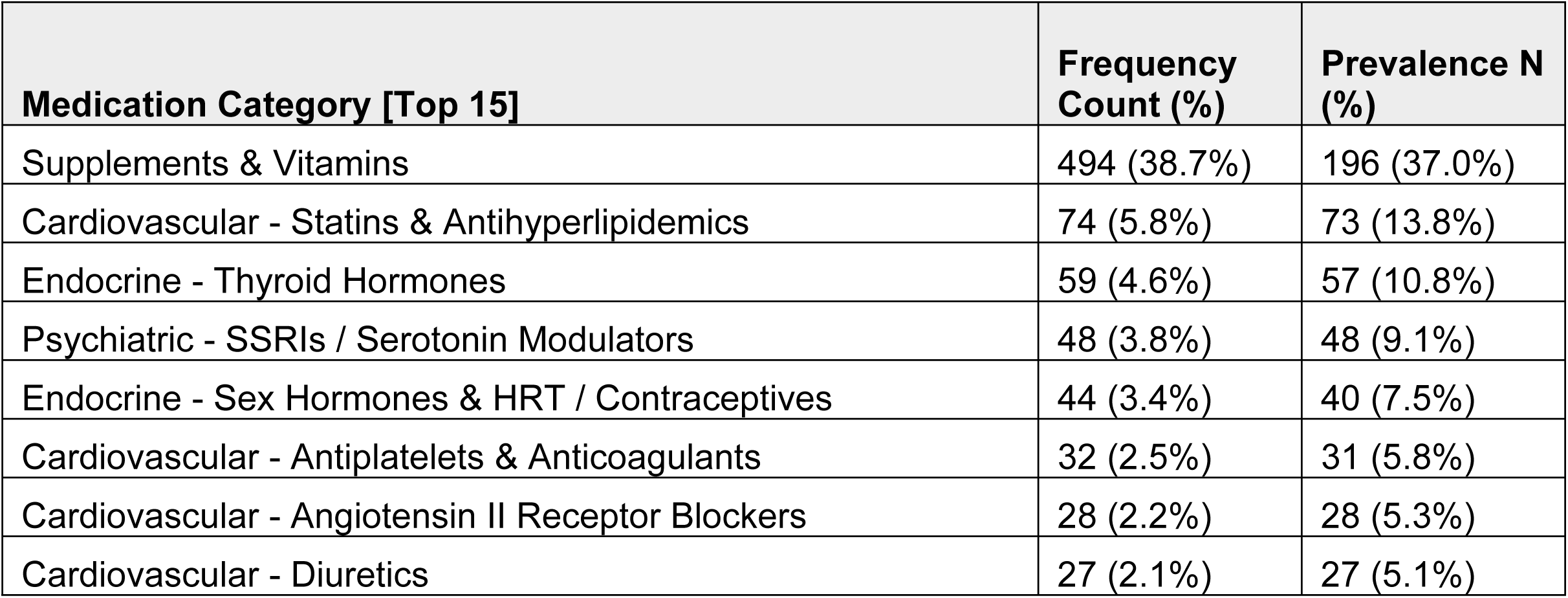
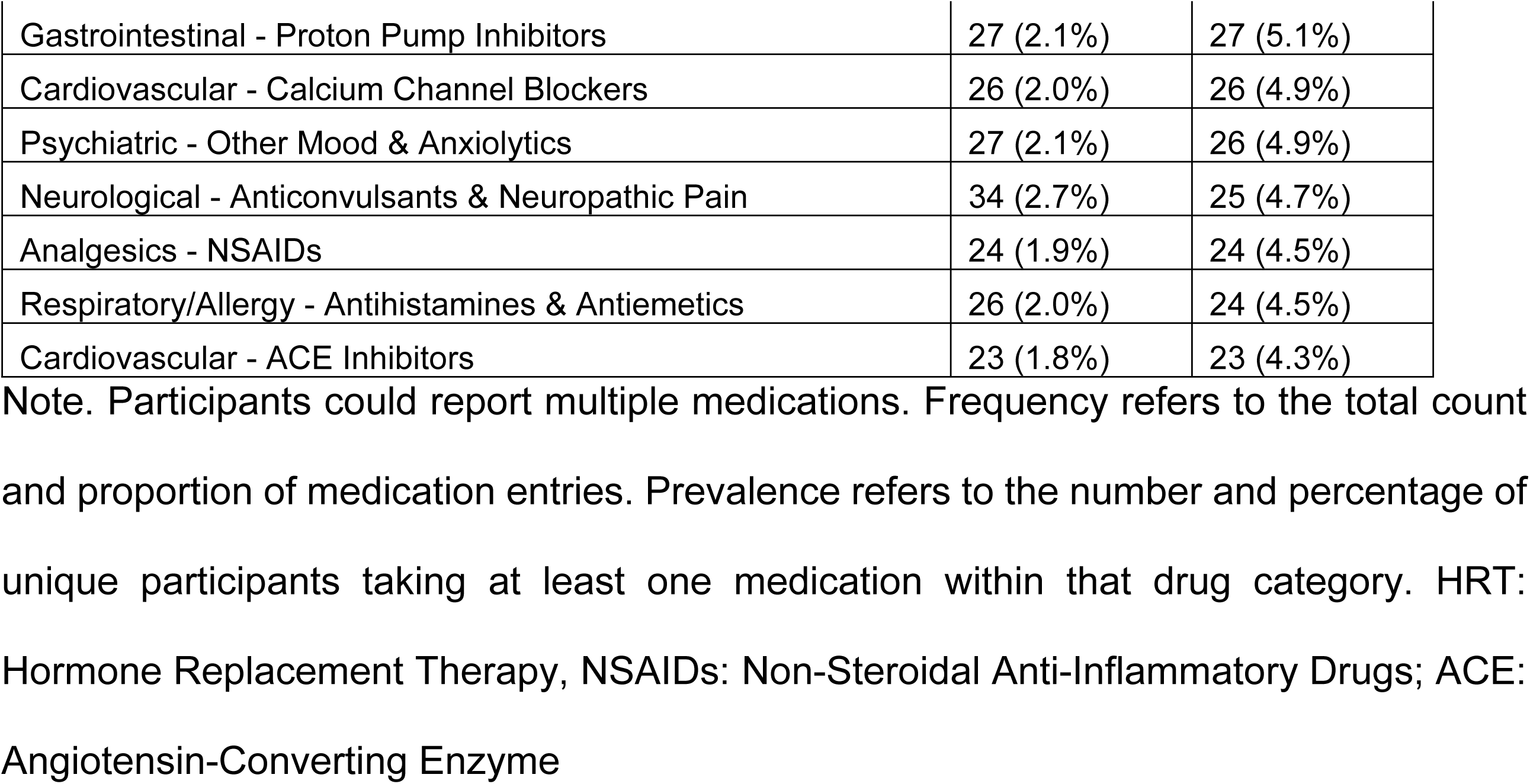
Top 15 Medication Categories Reported by Participants (N=358)

| **Medication Category [Top 15]** | **Frequency Count (%)** | **Prevalence N (%)** |
| --- | --- | --- |
| Supplements & Vitamins | 494 (38.7%) | 196 (37.0%) |
| Cardiovascular - Statins & Antihyperlipidemics | 74 (5.8%) | 73 (13.8%) |
| Endocrine - Thyroid Hormones | 59 (4.6%) | 57 (10.8%) |
| Psychiatric - SSRIs / Serotonin Modulators | 48 (3.8%) | 48 (9.1%) |
| Endocrine - Sex Hormones & HRT / Contraceptives | 44 (3.4%) | 40 (7.5%) |
| Cardiovascular - Antiplatelets & Anticoagulants | 32 (2.5%) | 31 (5.8%) |
| Cardiovascular - Angiotensin II Receptor Blockers | 28 (2.2%) | 28 (5.3%) |
| Cardiovascular - Diuretics | 27 (2.1%) | 27 (5.1%) |
| Gastrointestinal - Proton Pump Inhibitors | 27 (2.1%) | 27 (5.1%) |
| Cardiovascular - Calcium Channel Blockers | 26 (2.0%) | 26 (4.9%) |
| Psychiatric - Other Mood & Anxiolytics | 27 (2.1%) | 26 (4.9%) |
| Neurological - Anticonvulsants & Neuropathic Pain | 34 (2.7%) | 25 (4.7%) |
| Analgesics - NSAIDs | 24 (1.9%) | 24 (4.5%) |
| Respiratory/Allergy - Antihistamines & Antiemetics | 26 (2.0%) | 24 (4.5%) |
| Cardiovascular - ACE Inhibitors | 23 (1.8%) | 23 (4.3%) |

Note. Participants could report multiple medications. Frequency refers to the total count and proportion of medication entries. Prevalence refers to the number and percentage of unique participants taking at least one medication within that drug category. HRT: Hormone Replacement Therapy, NSAIDs: Non-Steroidal Anti-Inflammatory Drugs; ACE: Angiotensin-Converting Enzyme

### Dental History and Oral Hygiene

Dental history and oral hygiene habits are presented in Table 4. 56.8% of participants reported a history of cavities and 5.4% reported a history of gum disease. 91.6% reported having a regular dentist, 76% visit the dentist every 6 months, and 68.2% brush their teeth twice a day. Descriptive statistics for dietary and exercise habits are available in Supplementary Tables S3 and S4.

**Table 4:**
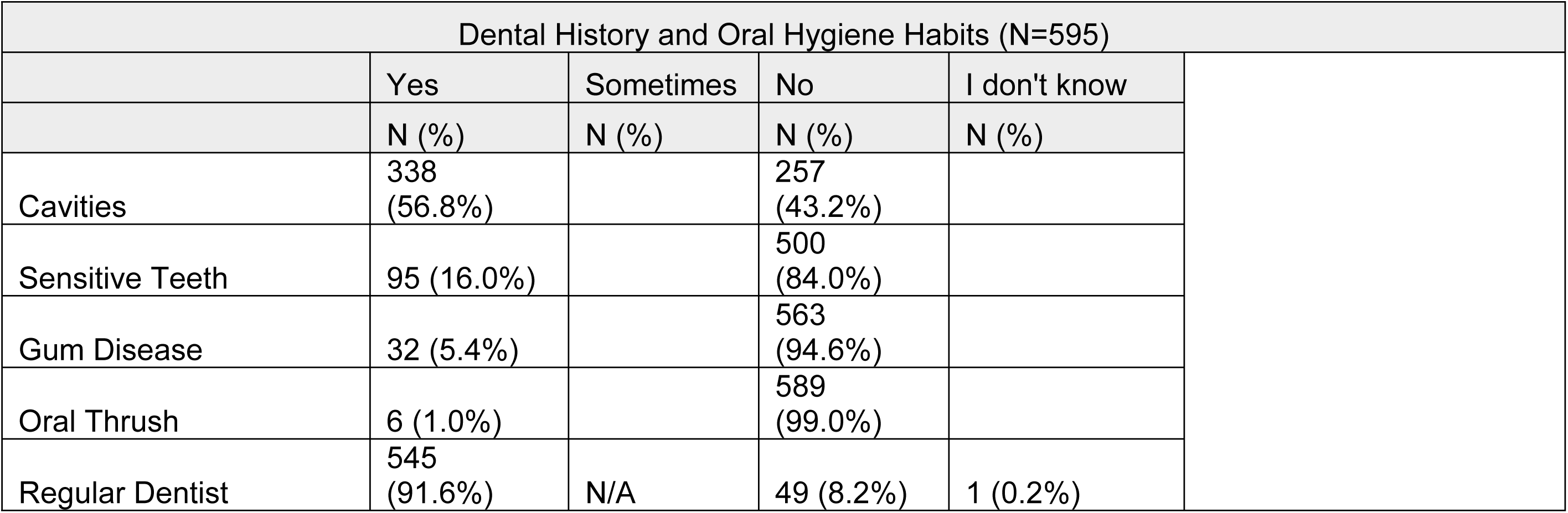
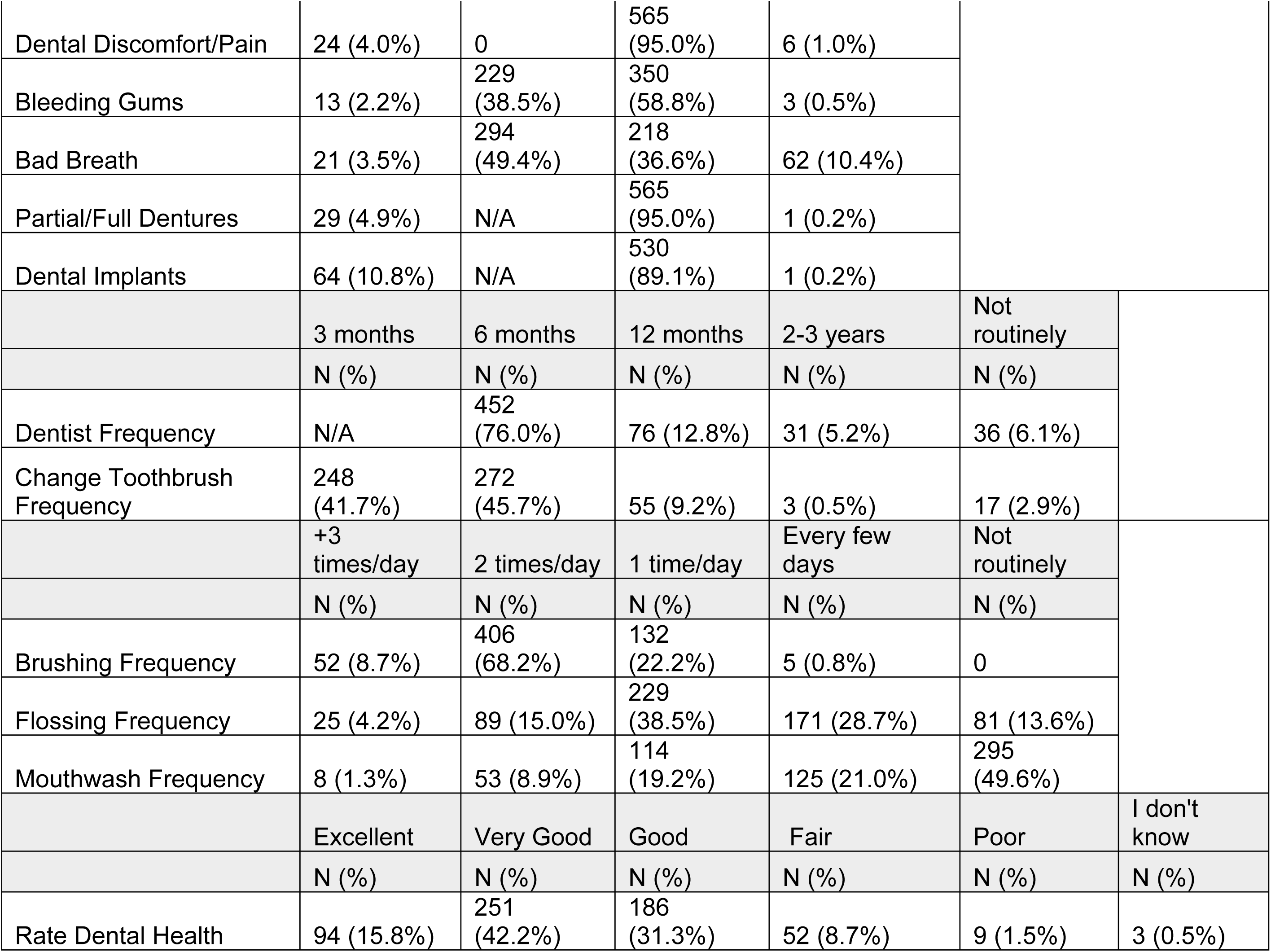
Dental History and Oral Hygiene Habits.

| Dental History and Oral Hygiene Habits (N=595) |  |  |  |  |
| --- | --- | --- | --- | --- |
|  | Yes | Sometimes | No | I don't know |
|  | N (%) | N (%) | N (%) | N (%) |
| Cavities | 338 (56.8%) |  | 257 (43.2%) |  |
| Sensitive Teeth | 95 (16.0%) |  | 500 (84.0%) |  |
| Gum Disease | 32 (5.4%) |  | 563 (94.6%) |  |
| Oral Thrush | 6 (1.0%) |  | 589 (99.0%) |  |
| Regular Dentist | 545 (91.6%) | N/A | 49 (8.2%) | 1 (0.2%) |

|  |  |  |  |  |  |  |
| --- | --- | --- | --- | --- | --- | --- |
| Dental Discomfort/Pain | 24 (4.0%) | 0 | 565 (95.0%) | 6 (1.0%) |  |  |
| Bleeding Gums | 13 (2.2%) | 229 (38.5%) | 350 (58.8%) | 3 (0.5%) |  |  |
| Bad Breath | 21 (3.5%) | 294 (49.4%) | 218 (36.6%) | 62 (10.4%) |  |  |
| Partial/Full Dentures | 29 (4.9%) | N/A | 565 (95.0%) | 1 (0.2%) |  |  |
| Dental Implants | 64 (10.8%) | N/A | 530 (89.1%) | 1 (0.2%) |  |  |
|  | 3 months | 6 months | 12 months | 2-3 years | Not routinely |  |
|  | N (%) | N (%) | N (%) | N (%) | N (%) |  |
| Dentist Frequency | N/A | 452 (76.0%) | 76 (12.8%) | 31 (5.2%) | 36 (6.1%) |  |
| Change Toothbrush Frequency | 248 (41.7%) | 272 (45.7%) | 55 (9.2%) | 3 (0.5%) | 17 (2.9%) |  |
|  | +3 times/day | 2 times/day | 1 time/day | Every few days | Not routinely |  |
|  | N (%) | N (%) | N (%) | N (%) | N (%) |  |
| Brushing Frequency | 52 (8.7%) | 406 (68.2%) | 132 (22.2%) | 5 (0.8%) | 0 |  |
| Flossing Frequency | 25 (4.2%) | 89 (15.0%) | 229 (38.5%) | 171 (28.7%) | 81 (13.6%) |  |
| Mouthwash Frequency | 8 (1.3%) | 53 (8.9%) | 114 (19.2%) | 125 (21.0%) | 295 (49.6%) |  |
|  | Excellent | Very Good | Good | Fair | Poor | I don't know |
|  | N (%) | N (%) | N (%) | N (%) | N (%) | N (%) |
| Rate Dental Health | 94 (15.8%) | 251 (42.2%) | 186 (31.3%) | 52 (8.7%) | 9 (1.5%) | 3 (0.5%) |

### Multi-omic Analysis

A total of 1,684 oral swab samples were collected from 595 subjects in seven batches over the 8-week study. This work presents comprehensive multi-omic data and multi-variate analyses generated from the baseline samples (metagenomics N=590; metabolomics N=595; immunological biomarkers N=595).

A principal component analysis of normalized metabolites with annotation levels of 1 and 2a showed comparable clustering across 570 metabolites except for only a few metabolites (Fig 1). Highly abundant metabolites found across samples were amino acids and derivatives, fatty acids and derivatives, nucleic acids and derivatives, sugars, tryptophan metabolites, acetaminophen derivatives, coumarin derivatives, and statin derivatives (Fig 1).

**Fig 1.**
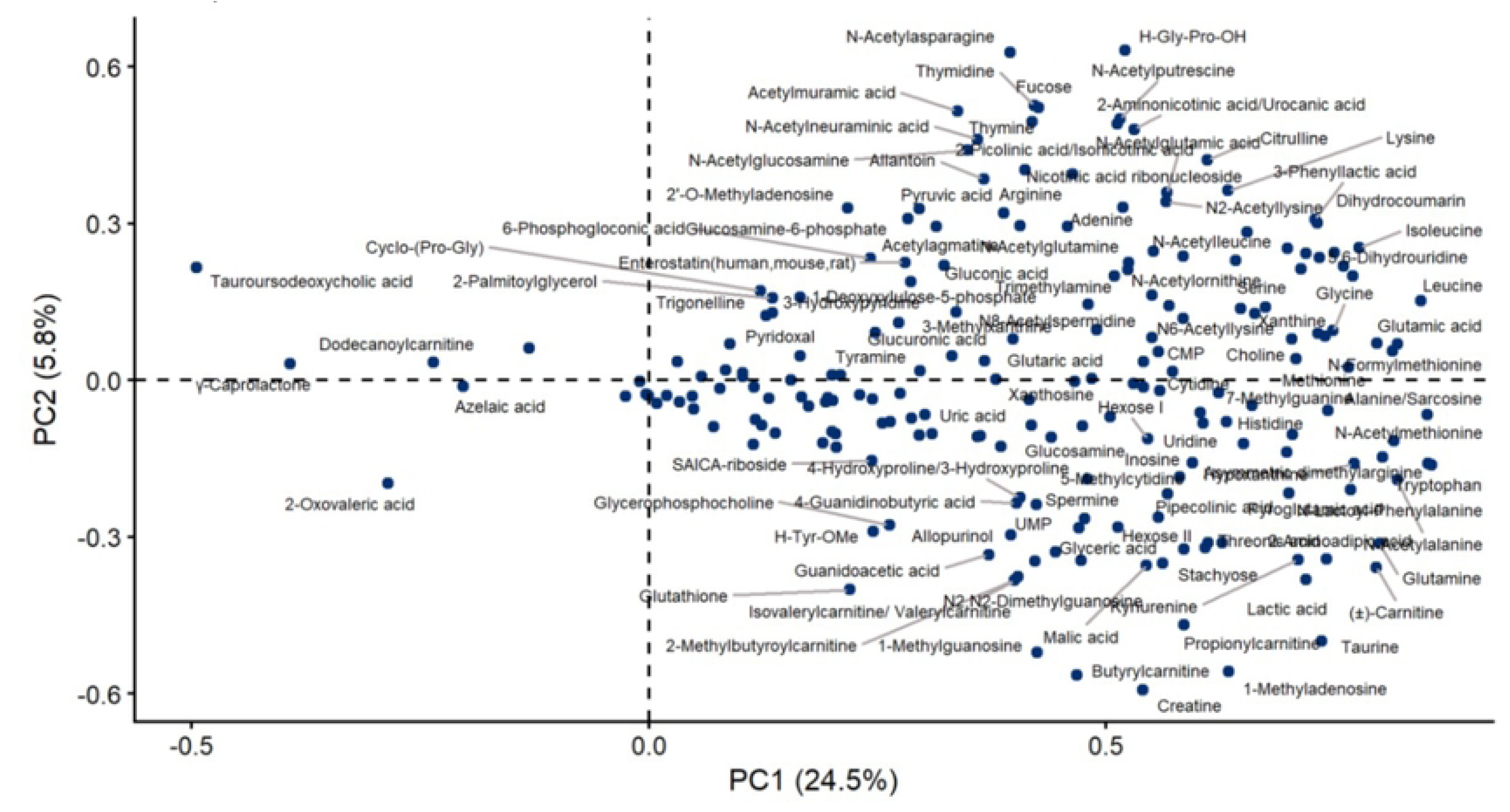
Loading plot from PCA model calculated on the normalized core data from the semipolar analysis. The x- and y-axis labels indicate the variance explained by the first two principal components. Data has been auto scaled and overlapping labels removed to improve readability.

### Oral Community Type (OCT) Analysis

Recovered sequences were assessed for relative abundance and queried against multiple microbiome databases comprising a total of 6,809 species. Following subtractive analyses of the environmental control swabs, 1,378 species were identified across all clinical samples with genus-level and species-level taxonomic analyses that differentially clustered into four distinct oral community types (OCT1, OCT2, OCT3, OCT4) using Dirichlet multinomial mixture modeling (Table 5). OCT 1 and OCT2 had the most shared metagenome profiles, while OCT4 had the most non-shared metagenome profiles. Fig 2A presents the top ten most relatively abundant Genera and species in each OCT group. On average, 193 microorganisms were detected per sample at a richness of 101 species, with the most predominant microbes belonging to the *Streptococcus* genera: *S. vestibularis, S. mitis, S. gordonii,* and *S. symci*. Other prevalent oral microbes identified included *Actinomyces* spp.*, Rothia* spp., and *Gemella* spp. OCT1, OCT2, and OCT3 contained some overlapping taxonomic groups but OCT4 had a notably higher abundance of *R. dentocariosa* compared to the other OCTs (Fig 2B).

**Table 5.** Oral Community Type (OCT) Summary.

|  | Proportional Size (%) | Homogeneity |
| --- | --- | --- |
| OCT1 | 34.5 | 40.4 |
| OCT2 | 27.7 | 53.6 |
| OCT3 | 23.2 | 31.5 |
| OCT4 | 14.6 | 14.6 |

**Fig 2.**
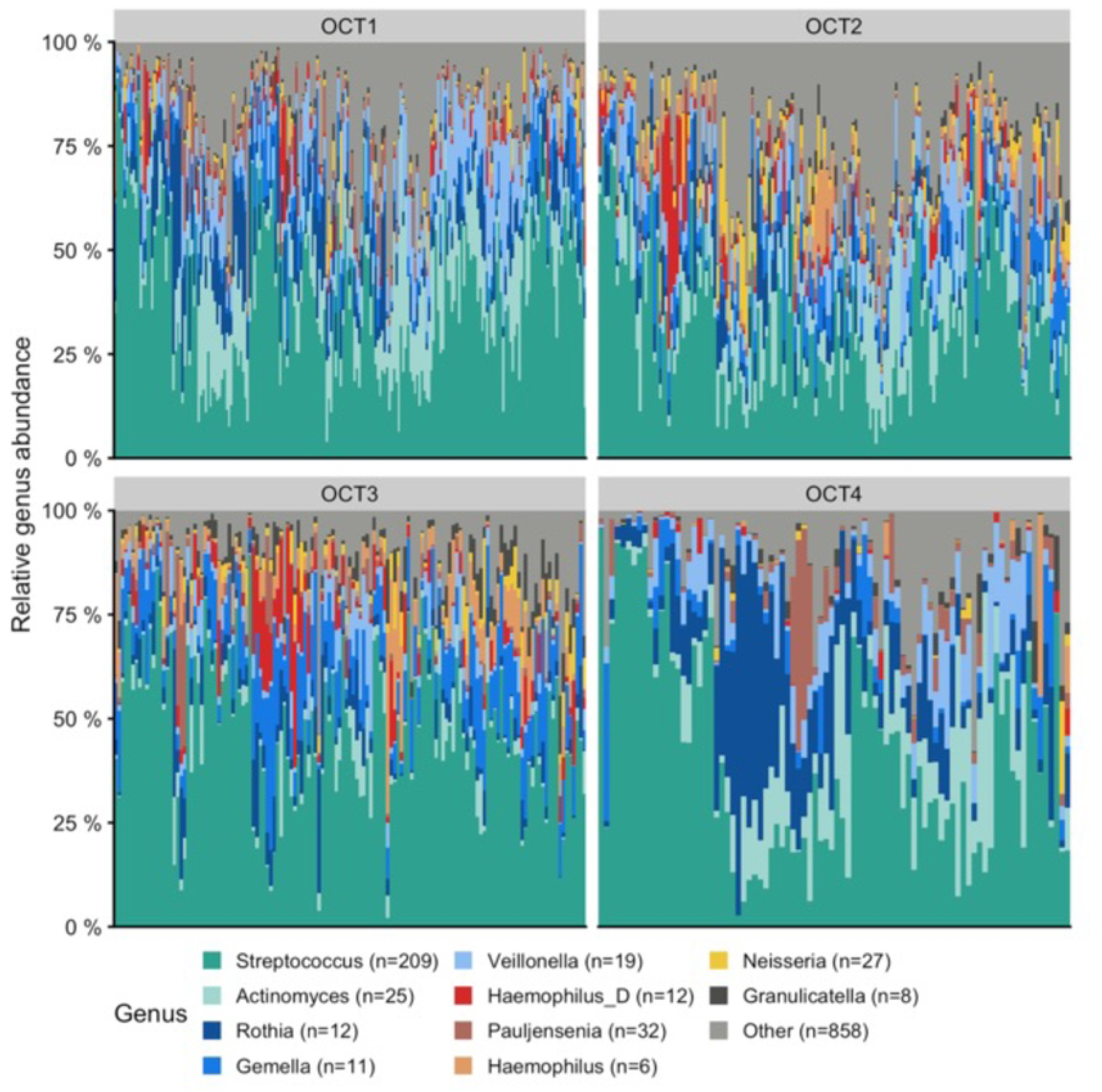
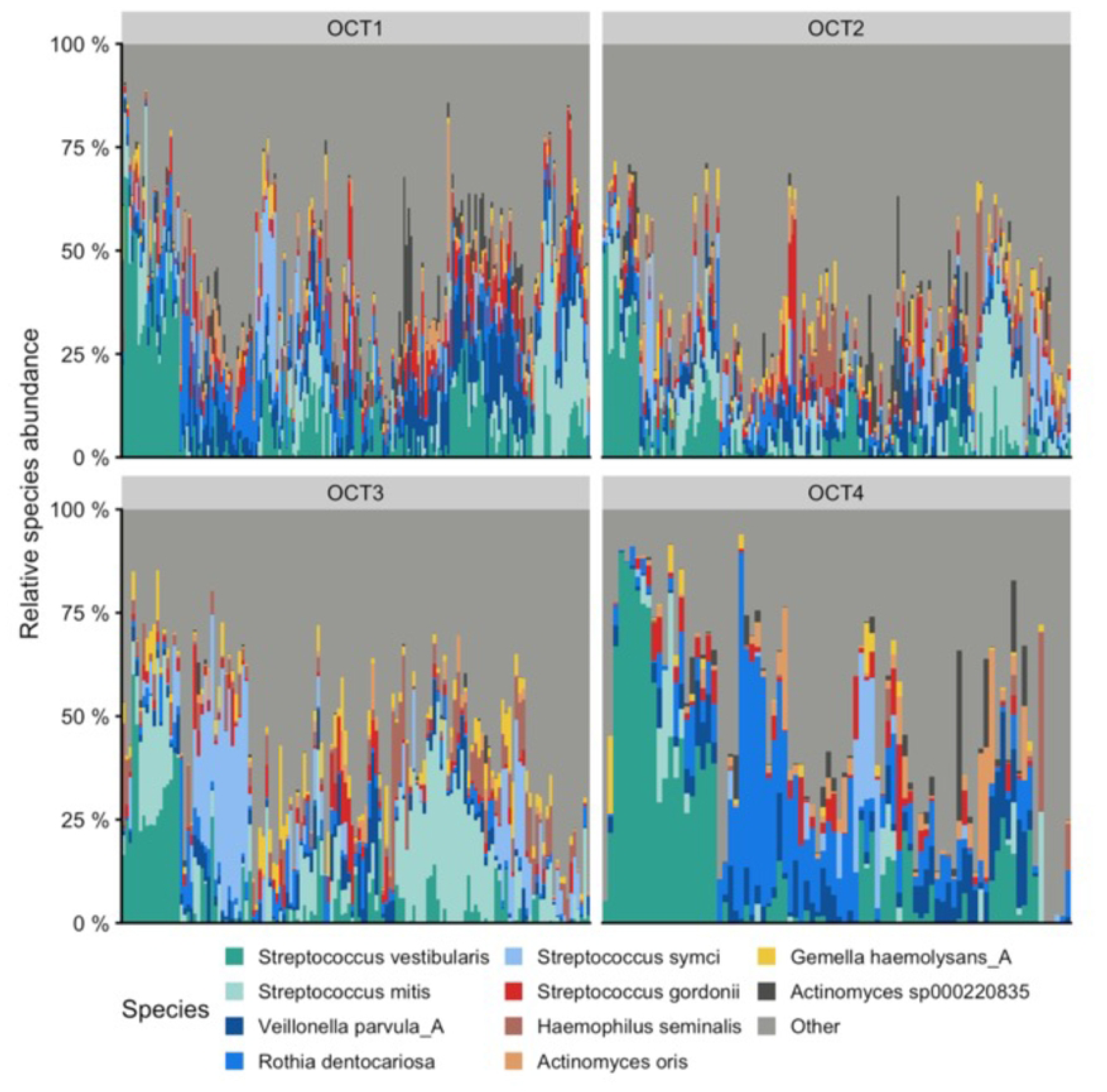
(A) Genus-level taxonomic overview split by OCT. For each sample, the bar plot indicates the relative abundance of the ten genera with the highest average abundance across all samples. Light grey (Other) indicates the total relative abundance of all other genera. (B) Species-level taxonomic overview split by OCT. For each sample, the bar plot indicates the relative abundance of the ten species with highest average abundance across all samples. Light gray (Other) indicates the total relative abundance of all other species.

Functional profiling was conducted using KEGG modules to characterize the metabolic capabilities and potential ecological function of these communities. The most abundant pathways across all samples are presented in Fig 3, which reflect fatty acid biosynthesis, carbohydrate metabolism, methanogenesis, aminoglycoside resistance, nucleic acid metabolism, and amino acid metabolism.

**Fig 3.**
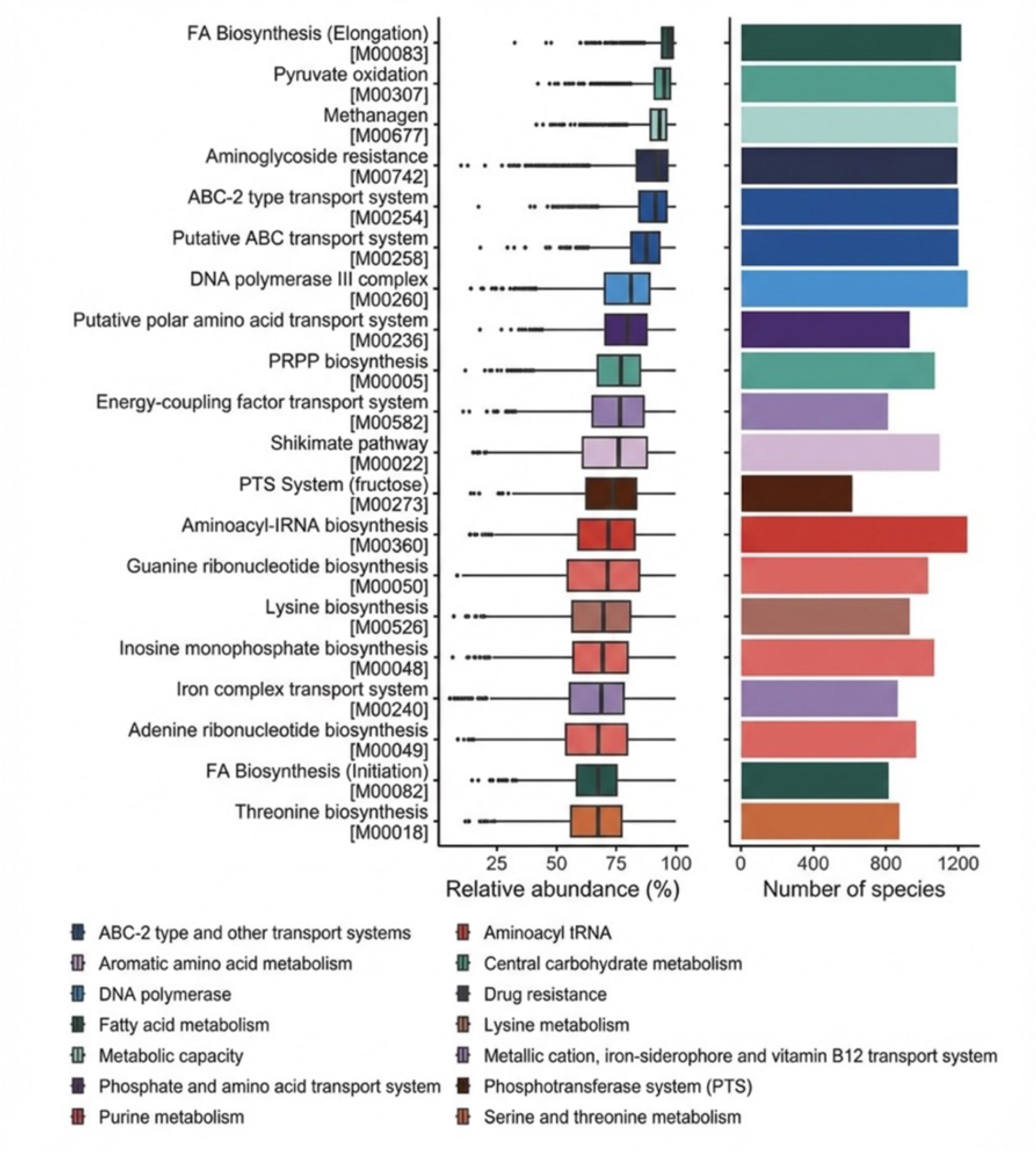
Boxplot and bar plot representing the top 12 KEGG annotation categories based on microbial genomic features associated with KEGG modules. The left panel shows the relative abundance (%) of each category, with boxplots ordered by decreasing median abundance, where each box represents the distribution of abundance values across samples. The right panel displays the count of microbial genomes containing the KEGG module for each category, with bars ordered to correspond with the left panel. Colors differentiate annotation categories, with the legend presented at the bottom for clarity.

Characterization of sex as an influence across OCTs is described in Table 6. OCT1 and OCT2 and OCT3 and OCT4 had a similar distribution of females-to-males, with a higher ratio of females-to-males in OCT3 and OCT4.

**Table 6.** Distribution of OCT Categories by Sex.

| OCT | Female N (%) | Male N (%) |
| --- | --- | --- |
| **1 (n = 203)** | 175 (86.2%) | 28 (13.8%) |
| **2 (n = 164)** | 139 (84.8%) | 25 (15.2%) |
| **3 (n = 137)** | 125 (91.2%) | 12 (8.8%) |
| **4 (n = 86)** | 78 (90.7%) | 8 (9.3%) |

The distribution of racial/ethnic background in each OCT is presented in Table 7. OCT1 and OCT4 had the highest percentages of Caucasian participants, with 72.9% and 79.1%, respectively. OCT3 was the most diverse, with 40.9 % being non-white, 24.1% of whom were Black. OCT2 comprised the second highest percentage of non-white participants at 34.7%.

**Table 7.** Distribution of OCT Categories by Race/Ethnicities.

| OCT | White or Caucasian N (%) | Black N (%) | Hispanic or Latino N (%) | Other/Multiracial N (%) |
| --- | --- | --- | --- | --- |
| **1 (n = 203)** | 148 (72.9%) | 28 (13.8%) | 8 (3.9%) | 19 (9.4%) |
| **2 (n = 164)** | 107 (65.2%) | 22 (13.4%) | 15 (9.1%) | 20 (12.2%) |
| **3 (n = 137)** | 81 (59.1%) | 33 (24.1%) | 11 (8.0%) | 12 (8.8%) |
| **4 (n = 86)** | 68 (79.1%) | 15 (17.4%) | 1 (1.2%) | 2 (2.3%) |

The distribution of age across OCTs is shown in Table 8. OCT2 and OCT3 were similar in average ages (51-52 years) and had similar age ranges. OCT4 had the highest average age (60 years) and the smallest age range (47 years).

**Table 8.** Oral Community Type Across Age.

| OCT | Mean Age (Years) | Age Range (Years) | 20-29 N (%) | 30-39 N (%) | 40-49 N (%) | 50-59 N (%) | 60-69 N (%) | 70-79 N (%) |
| --- | --- | --- | --- | --- | --- | --- | --- | --- |
| **1 (n = 203)** | 57.98 | 22-76 | 5 (2.5%) | 18 (8.9%) | 23 (11.3%) | 45 (22.2%) | 73 (36.0%) | 39 (19.2%) |
| **2 (n = 164)** | 52.4 | 21-78 | 14 (8.5%) | 29 (17.7%) | 29 (17.7%) | 26 (15.9%) | 51 (31.1%) | 15 (9.1%) |
| **3 (n = 137)** | 51.8 | 22-78 | 12 (8.8%) | 19 (13.9%) | 30 (21.9%) | 33 (24.1%) | 35 (25.5%) | 7 (5.1%) |
| **4 (n = 86)** | 60 | 28-75 | 1 (1.2%) | 9 (10.5%) | 12 (14.0%) | 16 (18.6%) | 38 (44.2%) | 10 (11.6%) |

To further characterize the beta-diversity within and between the OCTs at each time point, principal coordinates analysis of weighted UniFrac distances were used. Fig 4A shows how the OCTs cluster and their dispersion in beta-diversity space, respectively. The largest cluster, OCT1 overlapped the most with OCT2 and OCT4, while OCT3 showed separation on the first principal component basis with 35% of the variance assigned to this community signaling diverse species present amongst this cluster. Conversely, the smallest cluster, OCT4, was spread broadly over the first component, which fit with the low homogeneity within this cluster and was substantially different from OCT1 and OCT3 clusters at all time points. Fig 4B shows OCT dispersion in beta-diversity space with OCT1 and OCT2 having similar average distances to centroid. OCT3 had the shortest average distance to centroid and OCT4 had the farthest average distance to centroid in beta-diversity space.

**Fig 4.**
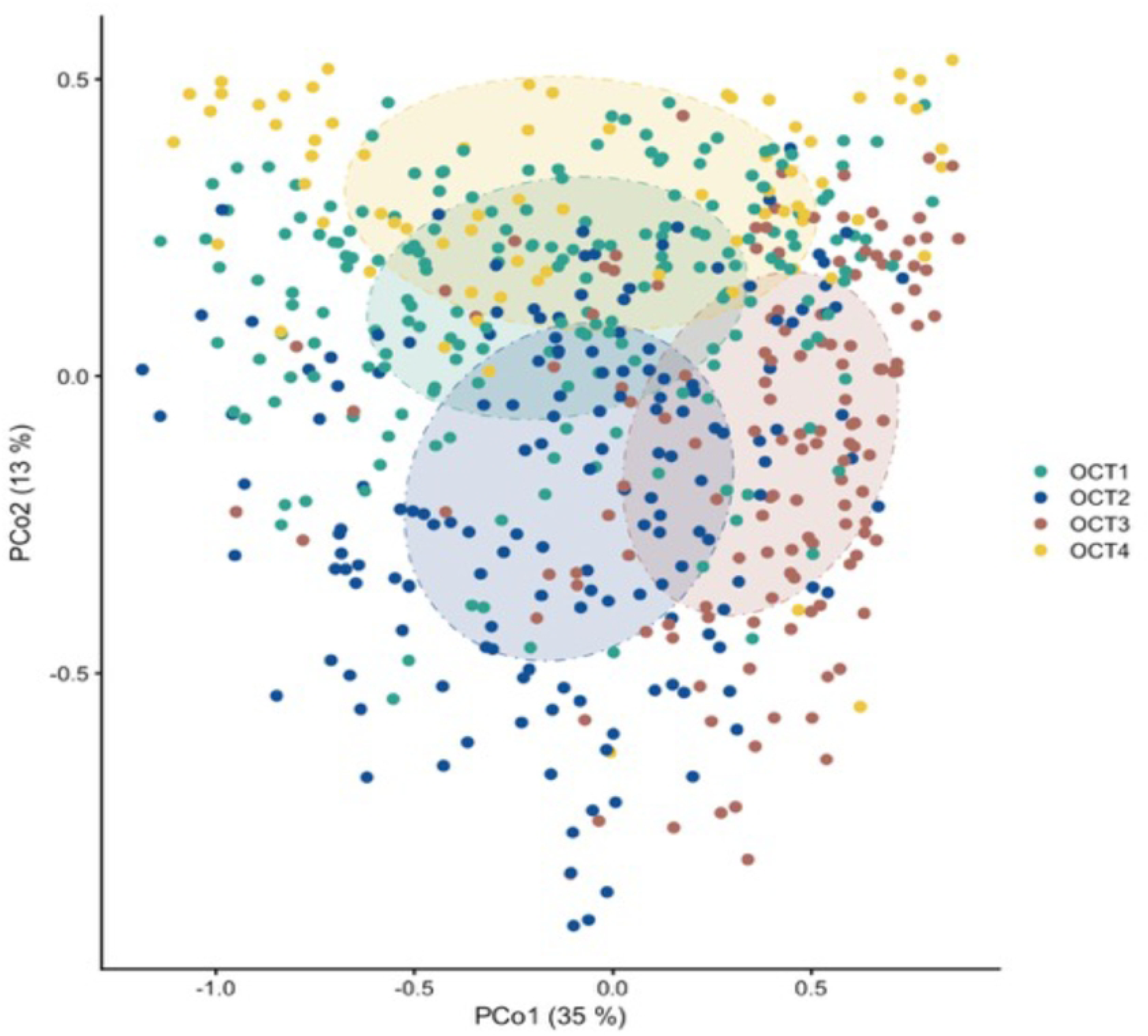
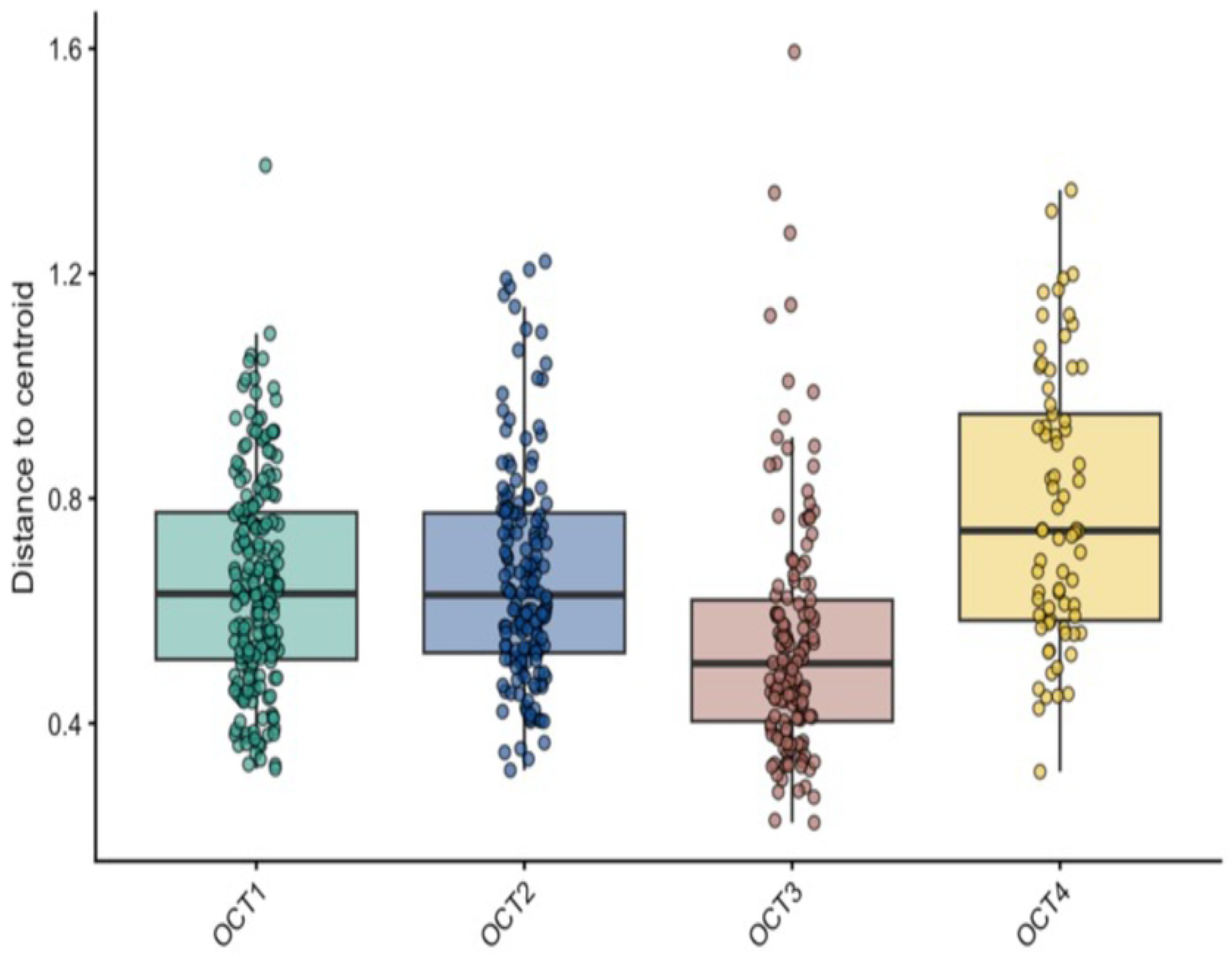
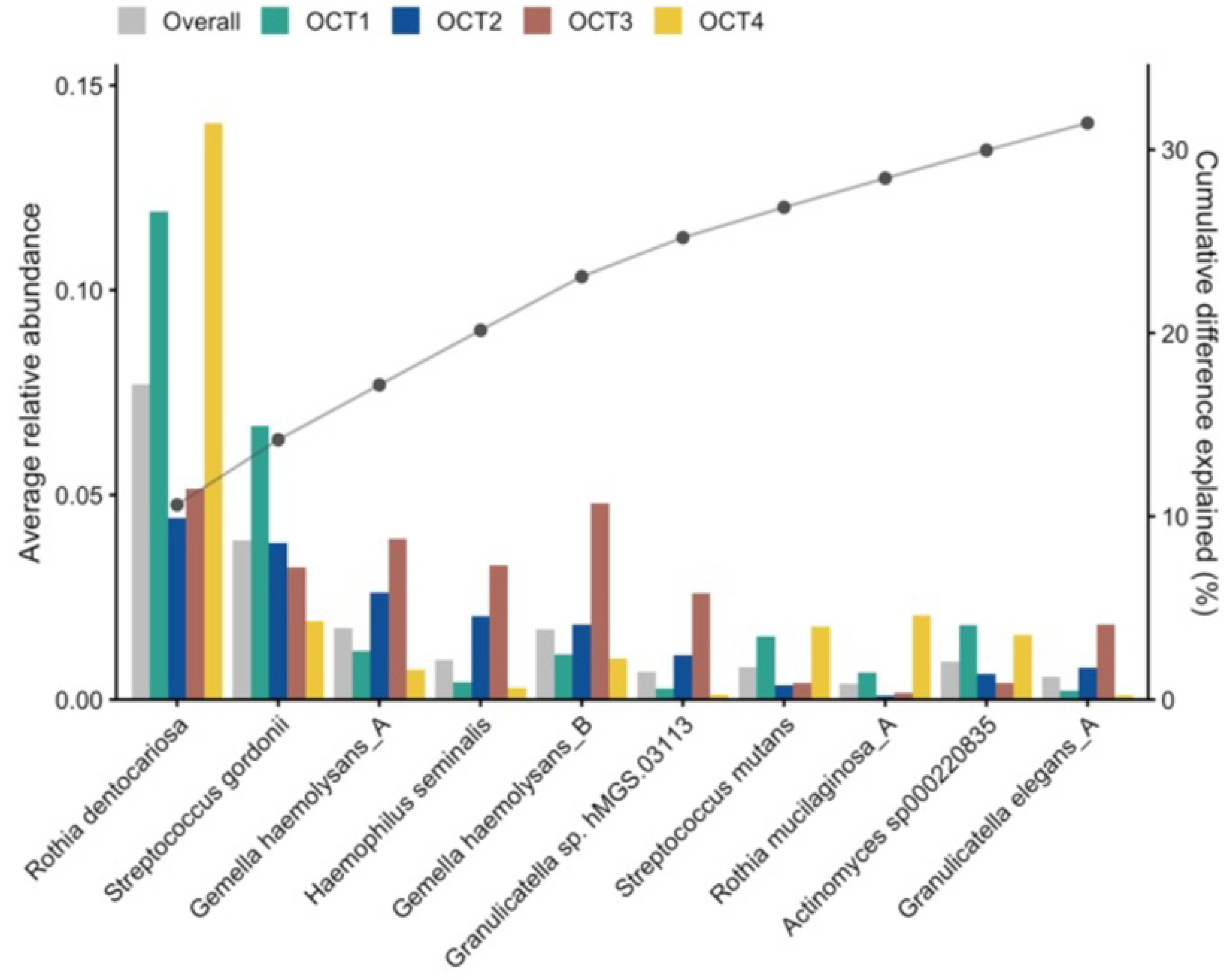
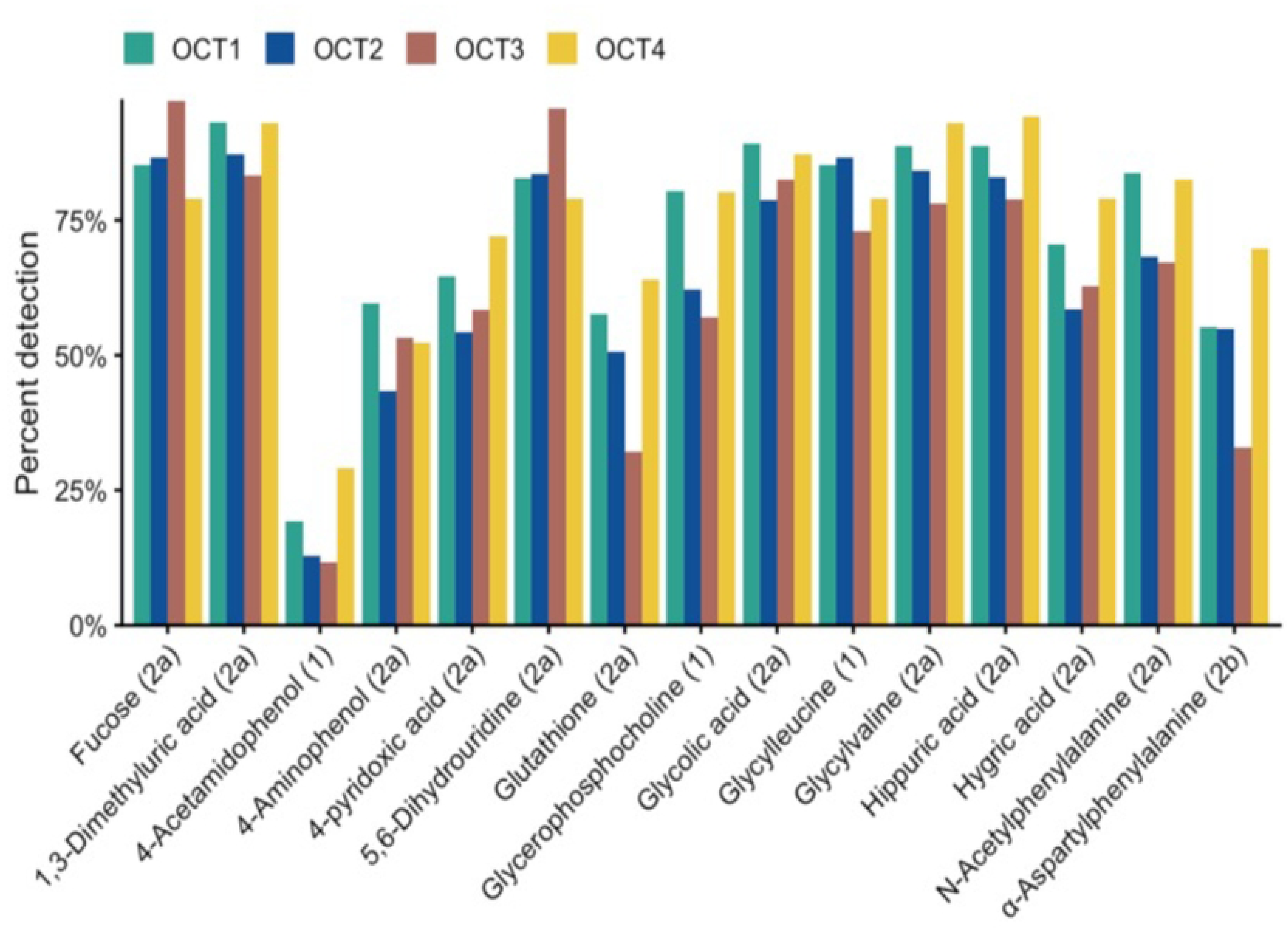
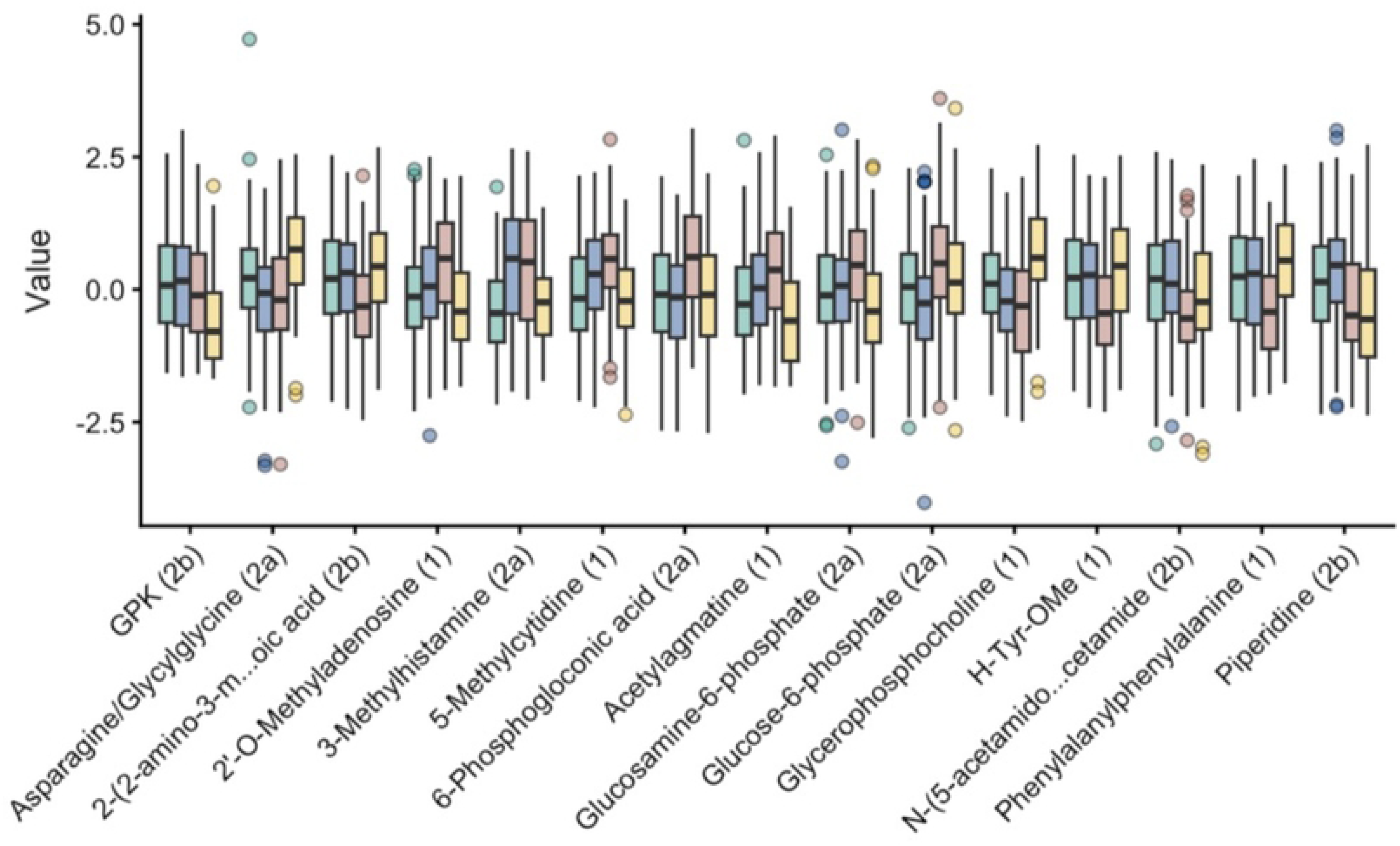
(A) Principal coordinates analysis (PCoA) based on weighted UniFrac distances among samples calculated based on the species abundances. Samples are colored by time-point. Ellipses represent the standard deviation of the samples within each time-point. The x- and y-axis labels indicate the microbial variance explained by the first two principal coordinates. (B) Homogeneity within each OCT. Distance from each time point to the centroid of its corresponding OCT in weighted UniFrac beta diversity space. (C) The ten species contributing the most to the oral community type clustering. The bars show the average relative abundances of each species overall (gray) and within each OCT. The line plot shows the cumulative difference between the overall average abundances and the average abundances in the OCTs that is explained by the species set. This value serves as an estimate of the variance explained by the species set. (D) Distribution of the 15 most associated detected compounds among the OCTs, as defined by the lowest FDR and belonging to annotation levels 1, 2a, or 2b. Compound detection is represented by its fraction within each OCT. (E) Distribution of the 15 most associated compound intensities among the OCTs as defined by the lowest FDR and belonging to annotation levels 1, 2a, or 2b. Compound intensities are represented by their median and interquartile ranges within each OCT. Whiskers extend to ± x the interquartile range and outlying samples are denoted by points.

Further analysis of the 10 most shared species across OCTs showed that the relative abundance of *Rothia dentocariosa* was a significant differentiating factor in OCT1 and OCT4 compared to OCT2 and OCT3 (Fig 4C). Additionally, OCT1 was also characterized by having a higher relative abundance of *Streptococcus gordonii* compared to the other community types. OCT3 was characterized by a higher abundance of *Gemella haemolysans, Haemophilus seminalis,* and *Granulicatella sp.* Together, these six bacterial species accounted for 25% of the differences across the population. Interestingly, the amount of *R. dentocariosa* in OCT2 was far lower in relative abundance compared to all other OCTs (Fig 4C).

Multinomial regression modeling identified a total of 420 metabolites with significantly different intensities that were associated with the OCTs, and 64 metabolites were differentially detected between OCTs at a level of FDR <0.1. Fig 4D presents the 15 most differentially detected metabolites between OCTs, including several sugar derivatives, organic acids, acetaminophen derivatives, dipeptides, and glutathione. Metabolite detection trends were similar between OCT1 and OCT4, while OCT3 and OCT4 showed the most divergence with respect to the 15 metabolites.

Analysis of the Intensity Values of top metabolites across OCTs differed from findings of Percent Detection analyses and are presented in Fig 4E. Similarly, di- and tripeptides and sugar metabolism intermediates were found, although the species differed. Notably, the tripeptide glycine-proline-lysine (GPK), the glucose metabolite phosphogloconic acid, neurotransmitter metabolites acetylagmatine and glycerophosphocholine, and the heterocyclic amine piperidine were annotated. The lowest intensity of GPK, acetylagmatine, glucosamine-6-phosphate, and piperidine was found in OCT4.OCT4 also had the highest intensity of Asp/GlyGly, and glycerophosphocholine compared to other OCTs. OCT2 and OCT3 shared the highest intensity of 3-methylhistamine. OCT2 had a higher intensity of piperidine and a lower intensity of glucose-6-phosphate (Fig 4E).

OCTs were also analyzed for levels of salivary Interleukin-6 (IL-6) and matrix metalloproteinases, MMP-8 and MMP-13 [31,32]. Interestingly, IL-6 levels were similar across OCTs (Fig 5A). MMP-8 levels were higher in concentration than IL-6 in OCT1, OCT2 and OCT4. OCT3 had the lowest concentration of MMP-8 across OCTs. MMP-13 was found to have the most significant association with OCT assignments (*P*-value <0.0001, Fig 5A).

**Fig 5.**
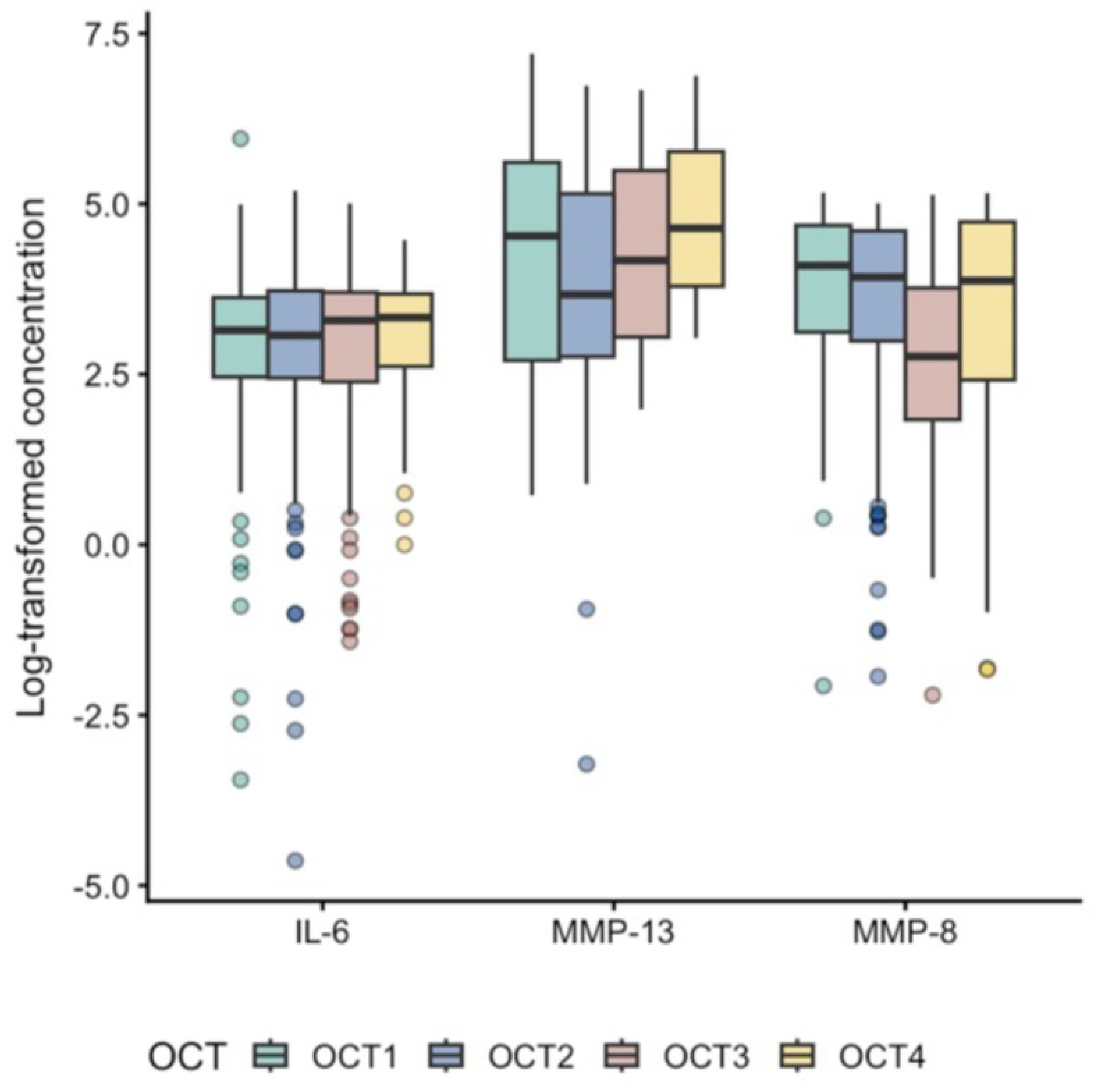
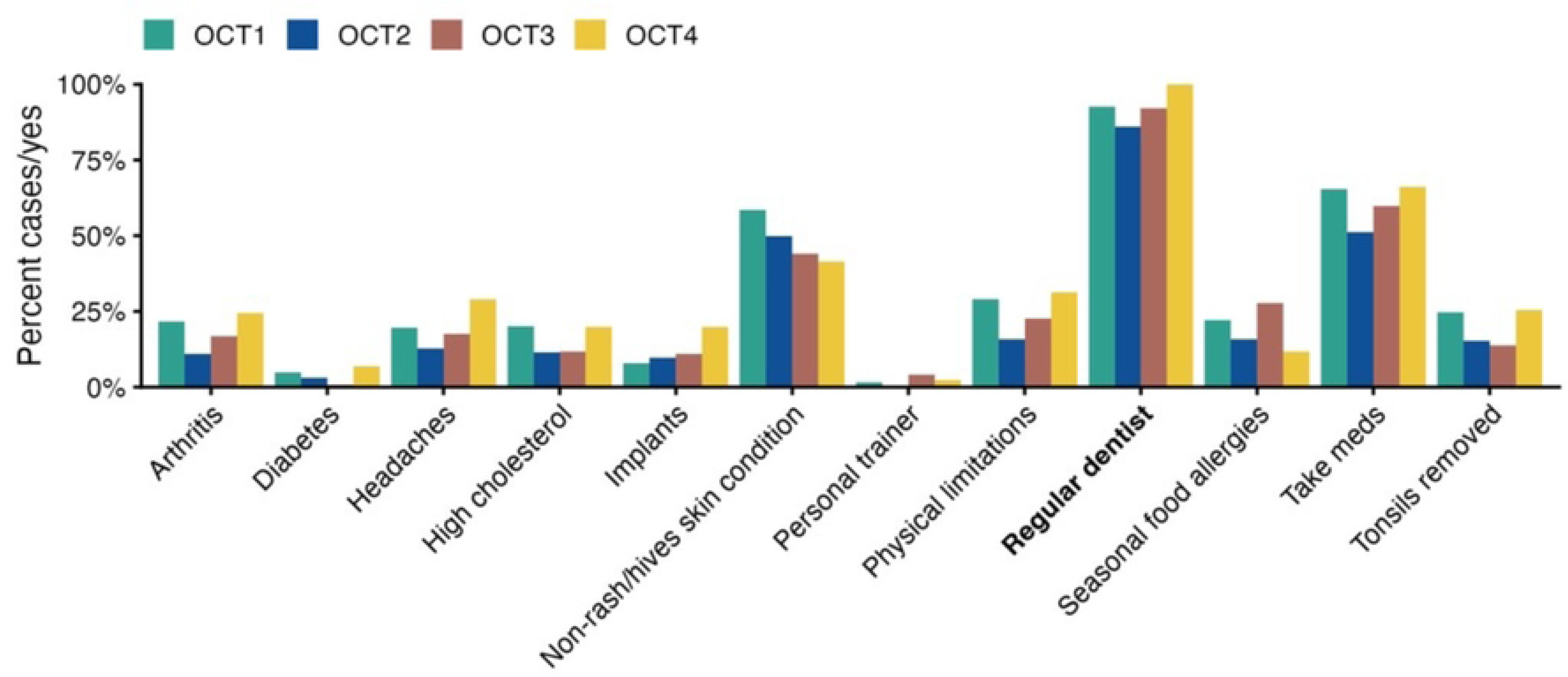
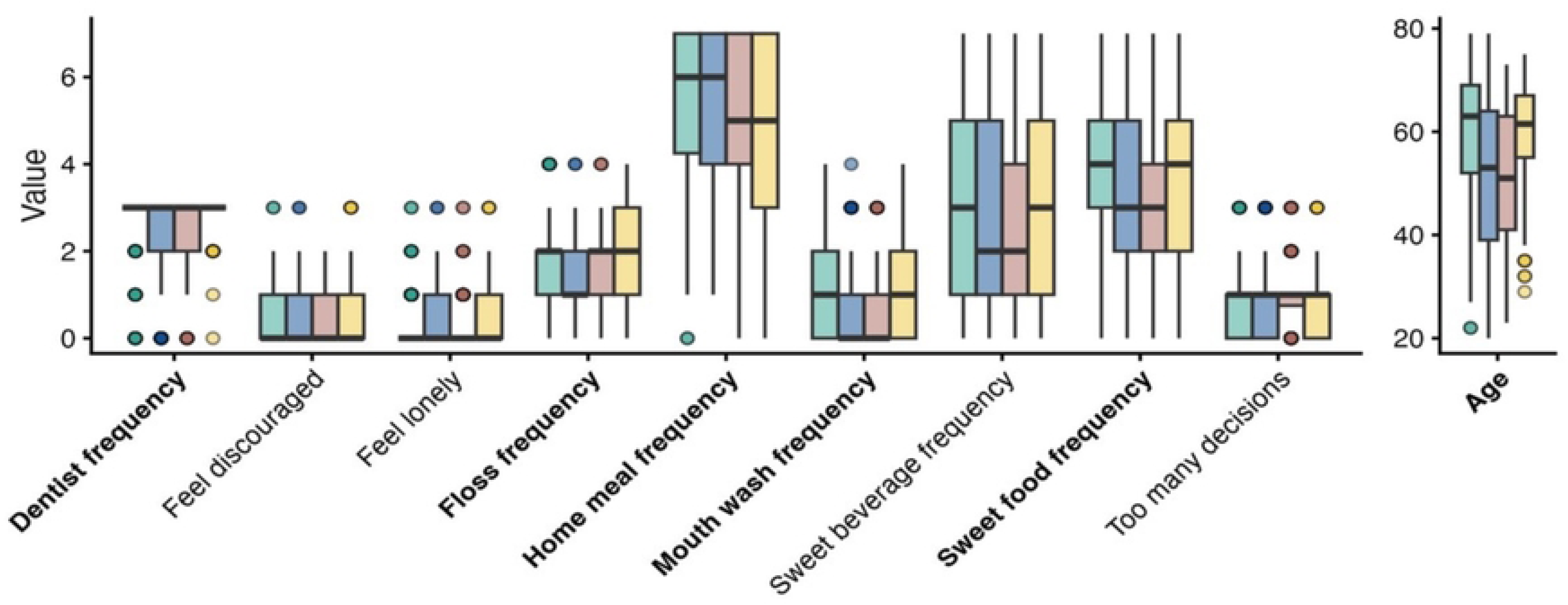
(A) Log transformed immune marker concentrations of samples with non-zero are represented by their median and interquartile ranges within each OCT. Whiskers extend to ± 1.5 x the interquartile range and outlying samples are denoted by points. (B) Distribution of questionnaire variables associated to the OCT assignments with a P-value of < 0.05. Binary variables are represented by the fraction of cases and “yes” responses within each OCT. (C) Distribution of questionnaire variables associated to the OCT assignments with a P-value of < 0.05. Ordinal variables are represented by their median and interquartile ranges within each OCT. Whiskers extend to ± 1.5 x the interquartile range and outlying samples are denoted by points. Variables found significantly associated at FDR < 0.1 are indicated in bold on the x-axis.

Of the binary variables, most participants visited a regular dentist, followed by medication usage (Fig 5B). OCT1 and OCT4 had similar distributions for many questionnaire variables apart from OCT4 reporting increased presence of dental implants and OCT1 reporting an increased history other skin conditions (non-rash, non-hives). OCT3 had the highest incidence of seasonal or food allergies and OCT2 had the lowest prevalence of participants that take medications, experience headaches, have arthritis, a personal trainer, and a regular dentist (Fig 5B).

Nine ordinal questionnaire features that significantly associated with OCT assignments at or below *P* < 0.05 and age are presented in Fig 5C. Dietary habits (frequency of consuming sweet foods and beverages, homemade meal prep), stress and emotion-related features were identified. Most significant, was the association of age across OCT assignments.

Eight questionnaire variables were significantly associated with OCT assignments based on a likelihood ratio test of a multinomial regression model (Table 6). OCT4 was highly associated with having a regular dentist, mouth washing frequency, floss frequency, and attending frequent dental visits. OCT1 was highly associated with Age, consumption of sweet food, and ethnicity. OCT2 was associated with eating home cooked meals frequently.

**Table 6.** Significant Associations Between OCTs and Questionnaire Features.

| Feature | FDR | Highest | Lowest |
| --- | --- | --- | --- |
| Age | 7e-10 | OCT1 | OCT3 |
| Has a Regular [non-specialty] Dentist | 0.0036 | OCT4 | OCT2 |
| Mouthwash Frequency | 0.0036 | OCT4 | OCT2 |
| Sweet Food Frequency | 0.0385 | OCT1 | OCT2 |
| Flossing Frequency | 0.0406 | OCT4 | OCT2 |
| Dentist Visit Frequency | 0.0438 | OCT4 | OCT2 |
| Home Meals Frequency | 0.0438 | OCT2 | OCT4 |
| Race/Ethnicity | 0.0900 | OCT1 | OCT3 |

The summary of the association results between microbiome taxon abundance, metabolite intensity, immune biomarkers and questionnaire from the linear regression with LinDA compositional bias correction features is presented in Table 7.

**Table 7.** Association of Microbiome and Metabolome Features with Participant Demographics, Oral Health Variables, and Immune Marker Detection

| Feature | Number of significant species/<br>Number of tested species | Number of significant compounds/<br>Number of tested compounds | Number of significant immune markers/<br>Number of tested immune markers |
| --- | --- | --- | --- |
| Diabetes | 12/290 | 11/664 | 0/3 |
| Age | 37/485 | 358/668 | 2/3 |
| Sex at Birth | 5/482 | 65/668 | 0/3 |
| Race/Ethnicity | 21/483 | 92/668 | 1/3 |
| Dental Visit Frequency | 12/485 | 26/668 | 0/3 |
| Rate Dental Health | 3/481 | 70/668 | 0/3 |
| Brushing Frequency | 5/485 | 3/668 | 1/3 |
| Flossing Frequency | 1/485 | 8/668 | 1/3 |
| Mouthwash Frequency | 12/485 | 27/668 | 0/3 |
| Changing Toothbrush Frequency | 0/485 | 0/668 | 0/3 |
| Bleeding Gums | 0/480 | 70/668 | 1/3 |
| IL-6 log concentration | 0/482 | 228/668 | N/A |
| MMP-8 log concentration | 76/485 | 558/668 | N/A |
| MMP-13 log concentration | 2/476 | 201/669 | N/A |
| IL-6 detection | 16/485 | 166/668 | N/A |
| MMP-13 detection | 1/485 | 448/668 | N/A |

Demographics such as age, race/ethnicity, and sex were significant with numerous metabolites and species. Conversely, oral hygiene habits such as visiting the dentist, tooth brushing frequency, flossing frequency, and using mouth wash were associated with fewer features. Notably, the frequency of changing one’s toothbrush was not associated with any baseline metabolites, species, or immune markers. MMP-8 concentration was significantly increased in participants that reported more frequent brushing, flossing, and bleeding gums.

## Discussion

This large public health study attempts to elucidate the connections between demographics, lifestyle, medical and -dental history, and oral hygiene metadata with untargeted metabolomics, deep shotgun metagenomic sequencing, and targeted immune marker measurements to gain a better understanding of the interrelationships that link the oral ecosystem to various systemic diseases, demographics, and lifestyle features. These data show statistically significant relationships between microbial species, gene pathways, metabolites, hygiene habits, medical conditions, and demographic features that correlate with dental health or self-reported bleeding gums.

Unexpectedly, 87.7% of study participants were female, much higher than the Minneapolis population (49.1%) [33]. However, this allowed for a more robust characterization of the female oral ecosystem. This is notable because females generally have higher rates of treated dental disease (fillings and restorations), increased prevalence of gingivitis (44.6% vs 27.3%), and decreased periodontitis (55.4% vs 72.7%) than males [34–39]. Ample evidence shows that hormones play a key role in oral health status throughout life, with an increasing risk of developing oral disease with age. The risk of developing periodontitis is higher after menopause [40]. Whether that is due to proactive versus avoidant healthcare behaviors, lifestyle, economics, or other socioeconomic and/or cultural drivers has yet to be determined.

The study population was moderately representative of the racial and ethnic demographics of Minneapolis, with Caucasians underrepresented (68.7% vs 79.8%) and Black participants more closely aligned with city averages (16.5% vs 18.8%) [33]. In accordance with the American Dental Association guidelines, 76.0% of participants reported regular dental visits (every 6 months), 68.2% brushed twice a day, and 41.7% replaced their toothbrush every 3 months. These results demonstrate high general compliance with preventive care, while also highlighting specific areas of unmet oral hygiene needs, such as 23% of participants that reported brushing once or every few days. Baseline questionnaires showed that 242 (40.7%) of the 595 study participants self-reported having or sometimes having bleeding gums, which correlates with prior studies [39] and contrasts in reported rates in other public health reports [41–43]. This is particularly interesting as studies have reported on the disparity between clinically observable bleeding gums and an over 50% lower propensity of those same patients to correctly self-report bleeding gums [41,44].

Notably, we found a high prevalence of mental health conditions, with 19.3% of participants reporting anxiety and 16.3% endorsing depression. Despite this, we observed a study mean PSQ-32 stress index of 0.25 which is slightly lower than the general population in Kocalevent et al., 2007 (0.30) [24]. However, in a post-hoc analysis of participants with anxiety and depression specifically, we observed an elevated PSQ-32 stress index of 0.30 in both groups which was significantly higher than the participants without anxiety or depression.

We also observed a high prevalence of other conditions such as arthritis (18.5%), high blood pressure (17.3%), high cholesterol (15.8%), and gastrointestinal issues such as acid reflux/GERD (11.8%) and irritable bowel syndrome (5%). Future studies are needed to explore potential mechanistic links between systemic health and variations in the oral microbiome and metabolome.

Analysis of the total counts of bacterial species and metabolites yielded surprising findings. Previous studies estimate that between 500 and 700 bacterial species are commonly found in the oral cavity, with 16s rRNA studies identifying 600 species [45,46]. This study identified 1,378 oral microbial species, which could reflect the increased sequencing depth, the use of two swabs used to capture sample at the tooth-gingival interface, or the study population.

A total of 1,513 metabolite compounds were detected after filtering, with 248 compounds characterized with high certainty, and 429 compounds matched based on mass, isotope pattern and reference counts. 836 compound signals remain unknown. Previous oral metabolome studies have cataloged 789 to 997 distinct core metabolites, and up to several thousand features without high confidence measures imparted [47–49].

The Oral Community Type (OCT) Analysis on the metagenomics sequencing revealed 4 OCTs with intriguing differences. OCT4 had the most heterogeneous speciation, characterized by a high abundance of *Rothia dentocariosa* and *Streptooccus mutans* and the lowest levels of *Streptococcus gordonii* and *Granulicatella spp.* The OCT4 cluster also reported more frequent and robust dental hygiene practices despite having the highest number of participants reporting dental implants, headaches, arthritis, and physical limitations. They also had the lowest incidence of self-reported bleeding gums although they had higher levels of MMP-8 and MMP-13.

Conversely, the most homogenous microbiome profile was present in OCT2, having the lowest abundance of *Rothia dentocariosa, Rothia mucilaginosa*, and *Streptococcus mutans* and the highest GPK, methylhistamine, and piperidine rates. This OCT also had the highest percentage of male participants, Hispanic and Asian participants, and reported less medication usage, and was less compliant with dental hygiene practices. Despite having the lowest MMP-13 and second lowest MMP-8 levels, OCT2 had the highest number of participants reporting bleeding gums.

OCT3 had the most distinct microbiome profile, characterized by a higher abundance of *Gemella spp*., *Granulicatella spp.,* and *Haemophilus spp.* compared to *Rothia spp.* and *Streptococcus spp.* Furthermore, OCT3 had the highest percentage of females, minority participants, and participants below the age of 50. They also reported less-frequent sweet food/beverage consumption, despite having the highest salivary glucose-6-phosphate, glucosamine-6-phosphate, and acetylagmatine level. OCT3 also had the second lowest rate of bleeding gums, and the lowest salivary MMP-8 concentration.

The OCT1 cluster had high levels of *Rothia dentocariosa* and *S. gordonii* approaching OCT4 levels, the highest MMP-8 and MMP-13 levels but had an average metabolomics profile. Behaviorally, they reported having a regular dentist yet attended dental visits infrequently. Interestingly, OCT1 had the lowest number of participants with dental implants, reported the second highest rate of bleeding gums, and the lowest methylhistamine levels.

Numerous studies show a strong correlation between MMP-8, MMP-13 and IL-6 with gingivitis and/or periodontitis [42,50–52]. It was therefore unexpected that the highest concentrations of MMP-8 and MMP-13 were found in the OCT with the lowest incidence of bleeding gums (OCT4). However, we did see significant associations between MMP-8 increased frequency of brushing, flossing, and bleeding gums across the entire sample. This positive association may indicate that mechanical irritation from brushing and flossing teeth may inflame periodontal tissues which can trigger a localized host immune response, resulting in active MMP-8 release.

Overall, IL-6 levels were moderate across all OCTs in the study population and did not correlate with MMP-8 and MMP-13 levels or self-reported bleeding gums, which is counter to previous work showing that IL-6 and MMP-8 are frequently identified together as promising salivary biomarkers for the early diagnosis and monitoring of gingival inflammation [42,50,51,53–55].

Interestingly, in this study, where dental disease was not used as an enrollment criterion, there were not any microbial species that correlated significantly with bleeding gums. In studies where clinically scored bleeding gums and/or periodontitis were used as enrollment criteria, significant correlation has been reported between disease severity and specific microbial species [56,57]. Of particular interest, 70 metabolites strongly correlated with bleeding gums, which builds upon previous work in gingivitis samples showing that metabolite shifts occur throughout the disease course and are statistically correlative [47].

On the contrary, microbial species were found to be significantly correlated with the abundance of MMP-8 and MMP-13, but not IL-6. The results in this study confirm previous work showing that microbial species are positively correlated with MMP-8 and MMP-13 levels [52–54].

Unexpectedly, the number of significant metabolites exceeded the number of significant microbial species with respect to questionnaire features and immune markers; the exception being frequency of toothbrush replacement, which had no significant association with microbial species or metabolites. In line with emerging reports, metabolites strongly correlated with immune marker levels, such as IL-6 (228 metabolites), MMP-8 (558 metabolites), and MMP-13 (201 metabolites) [55].

There were 12 significant microbial species and 11 significant metabolites that associated with study participants with a history of type 2 diabetes. Further clinical and mechanistic studies are needed to better understand the mechanisms underlying these relationships.

Interestingly, age and race/ethnicity correlated more strongly to specific microbial species and metabolites than hygiene habits (frequency of brushing, flossing, mouthwash use) or dental visit frequency. This emphasizes the importance of adjusting for age and race/ethnicity as confounding factors in future oral microbiome studies.

Since the late 1800s, significant advances in oral hygiene product development and their optimized frequency of use have been made, including antibiotics, antiseptic mouth rinses, nylon dental floss, and the electric toothbrush. These interventions focus on mechanical biofilm removal and eliminating pathogenic microorganisms with the goal of improved clinical outcomes [58]. Data from this study showing that 18 microbial species, 38 metabolites, and 1 immune marker (MMP-8) were shown to be significantly related to brushing, flossing and/or mouthwash was compelling. *Gemella haemolysans, Gemella haemolysans_B, Granulicatella sp. hMGS.03113*, 2-Picolinic acid/Isonicotinic acid, and Nicotinic acid ribonucleoside were associated with both mouthwash use and brushing frequency. Flossing frequency and mouthwash usage were both associated with 1-Methyl-3-(2-thiazolyl)-1H-indole. Also intriguing was the larger number of microbial species and metabolites that strongly associated with age, race/ethnicity, and immune markers. Future research is required to further elucidate the mechanisms underlying these associations.

Additionally, more work is planned to elucidate the associations between metabolites, microbial species and questionnaire features such as dental-medical history and medications.

Initial findings from this large multi-omic dataset and connected metadata provide a foundation to inform future “precision public health” research initiatives and support the development of more effective strategies for disease prevention and management. Further investigation of this dataset is needed to advance public health policy and clinical practice regarding oral-systemic disease across both the general population and specific subpopulations.

## Data Availability

The minimal data set is available at Dryad via 10.5061/dryad.3bk3j9m1s

## Acknowledgements

Delta Dental of Minnesota Foundation, Comprehensive Research Group, Cmbio, Spencer Wood, D.D.S., Raphael Eiesenhofer.

## Supporting Information Captions

**Supp. 1.**
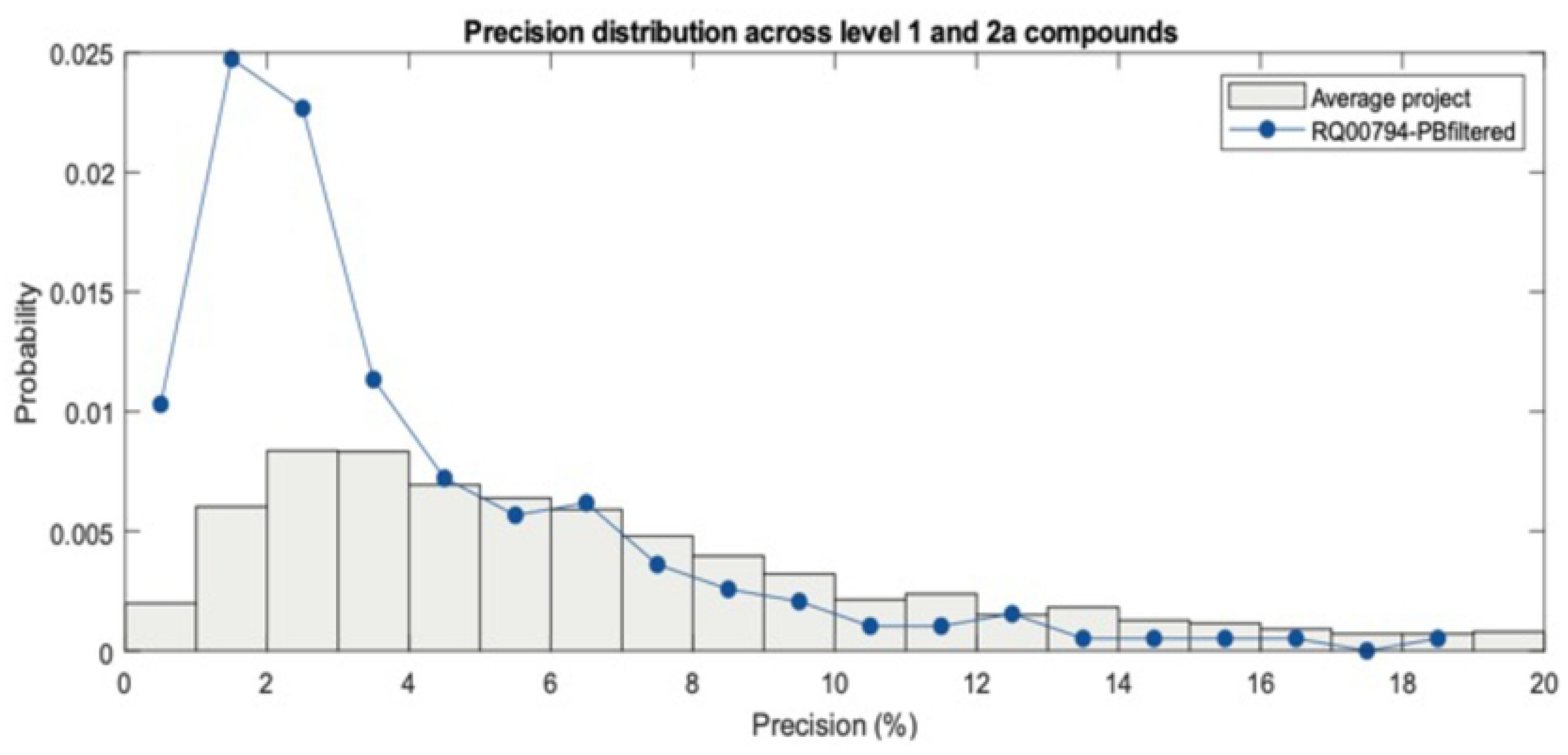
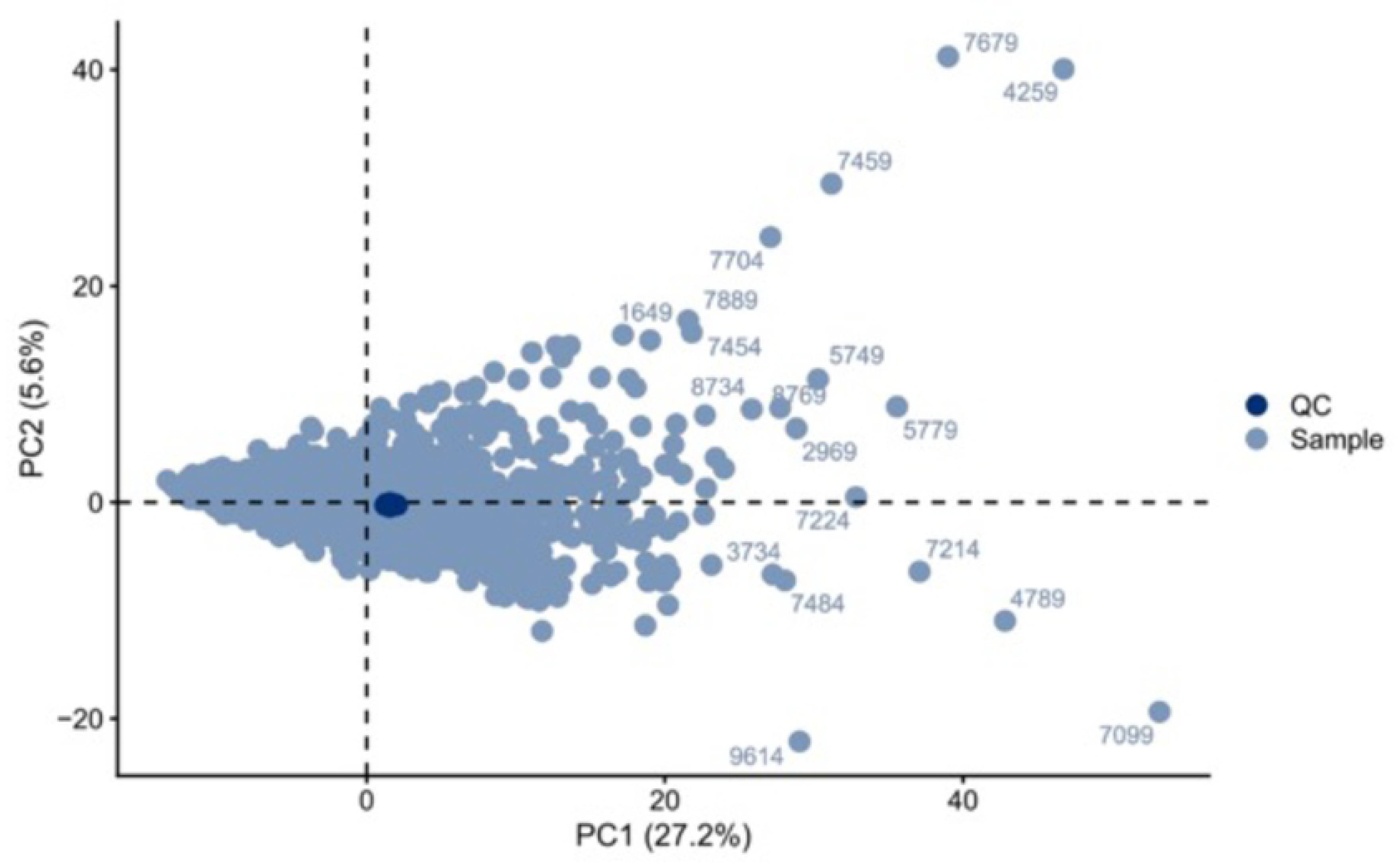
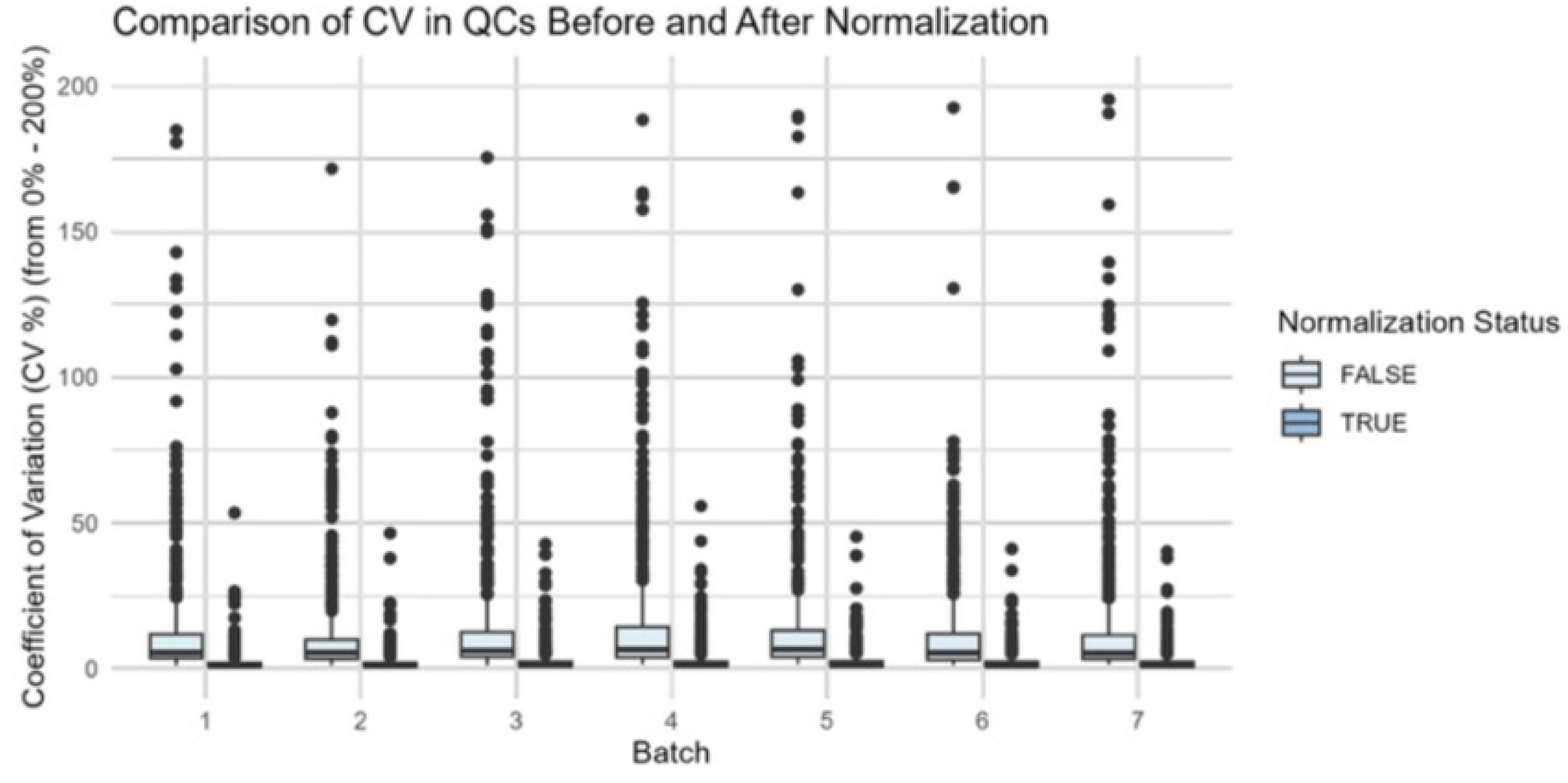
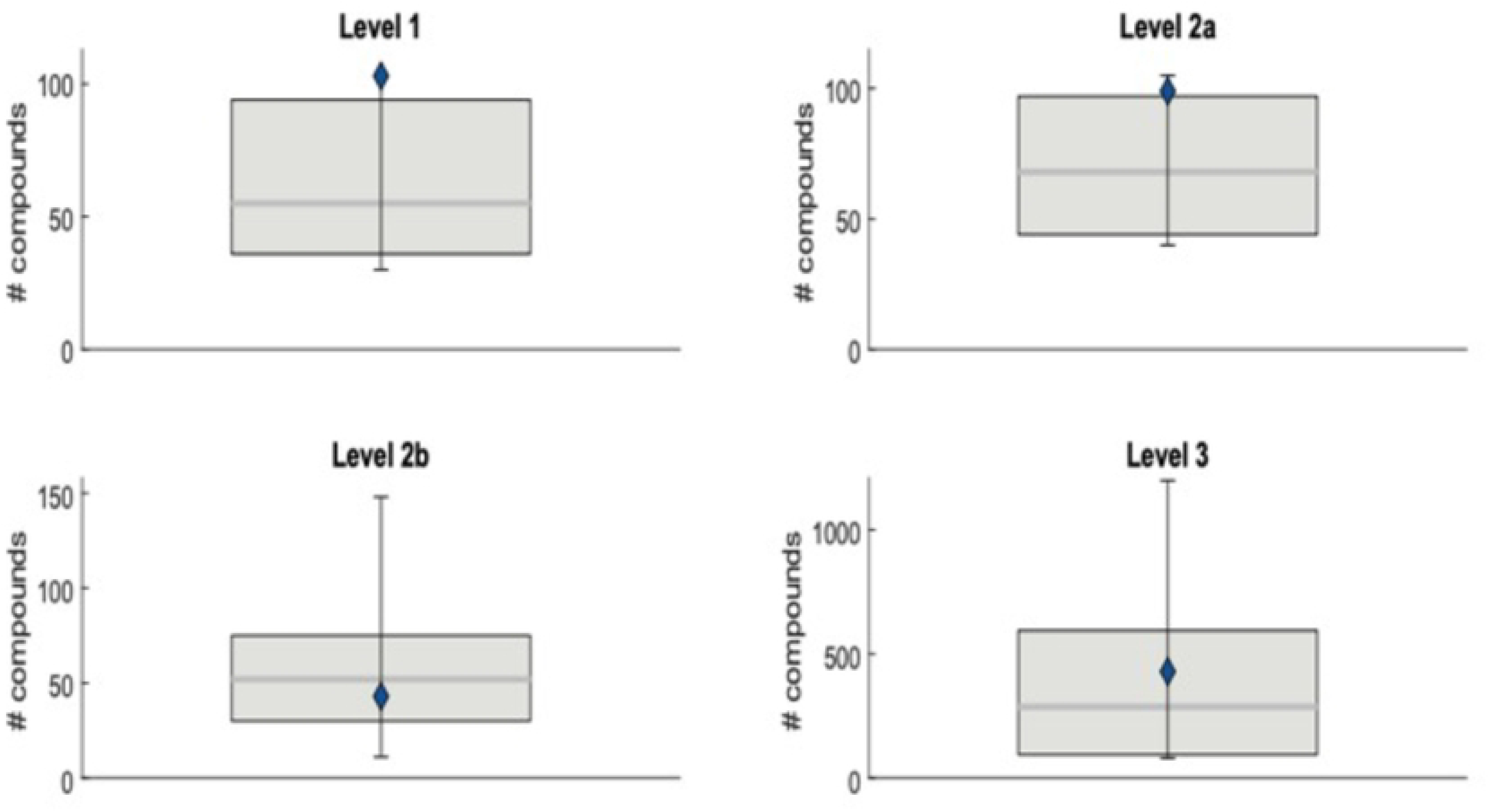
(A) Histogram of precision distribution across compounds on levels 1 and 2a. The blue points show the precision for this study, whereas the bars show the precision in an average study with a similar matrix (human fluids). A lower precision percentage is considered better. (B) Score plot of PC2 over PC1 from the PCA model calculated on the core data from the semi-polar analysis. The x- and y-axis labels indicate the variance explained by the first two principal components, and the colors separate the internal QC samples from the client samples. Edge samples have been labelled with client identifiers (barcodes). Data has been autoscaled. (C) Coefficient of variation for the 570 most intense compounds in the QCs, before and after SERRF normalization, across all batches. A decreased CV after normalization indicates a successful removal of analytical variance. (D) Box plots indicating the number of annotated compounds on each annotation level. The blue point shows the number for this study, whereas the boxplot shows the range of annotated compounds in other studies with a similar matrix.

**Suppl. 2.**
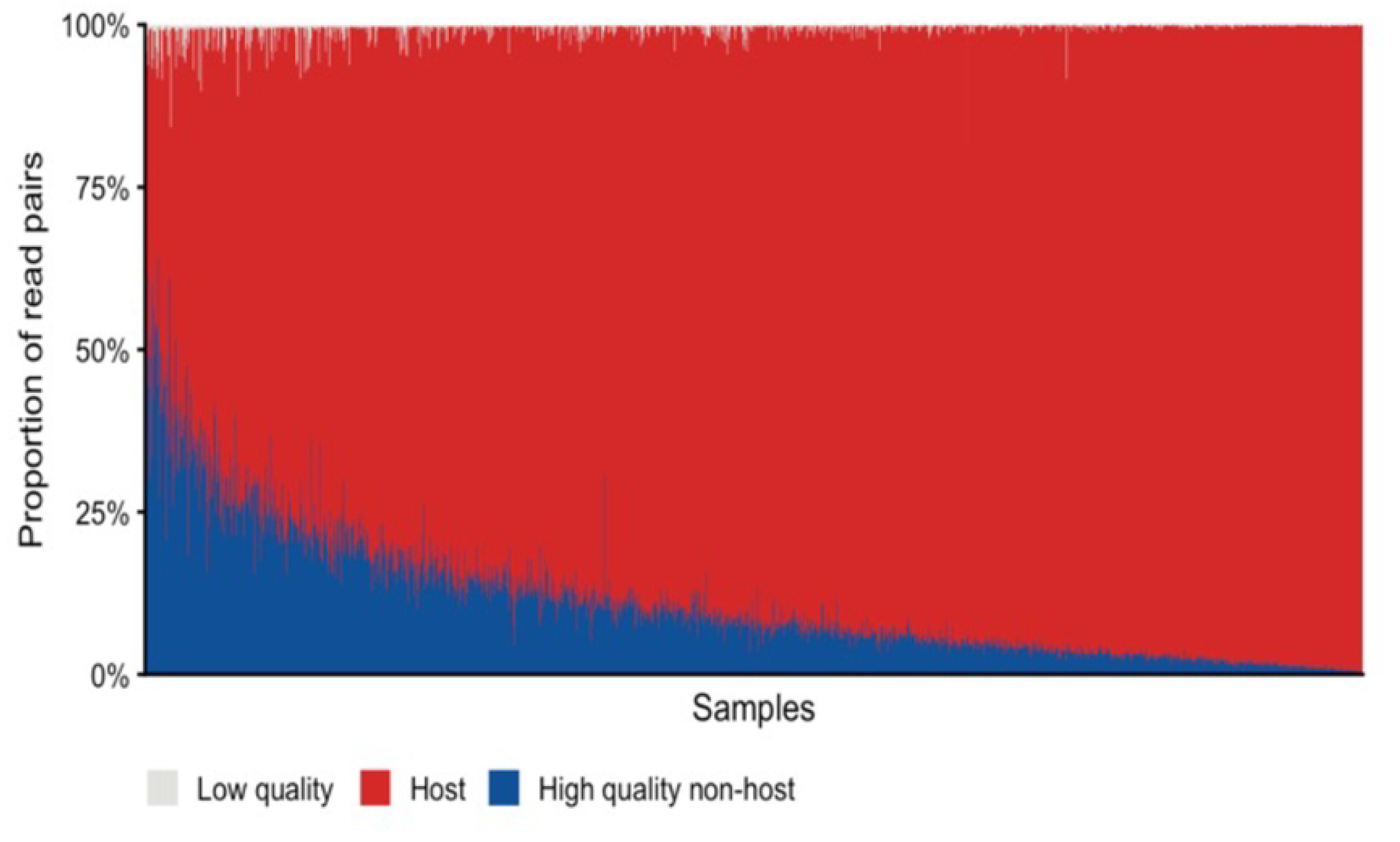
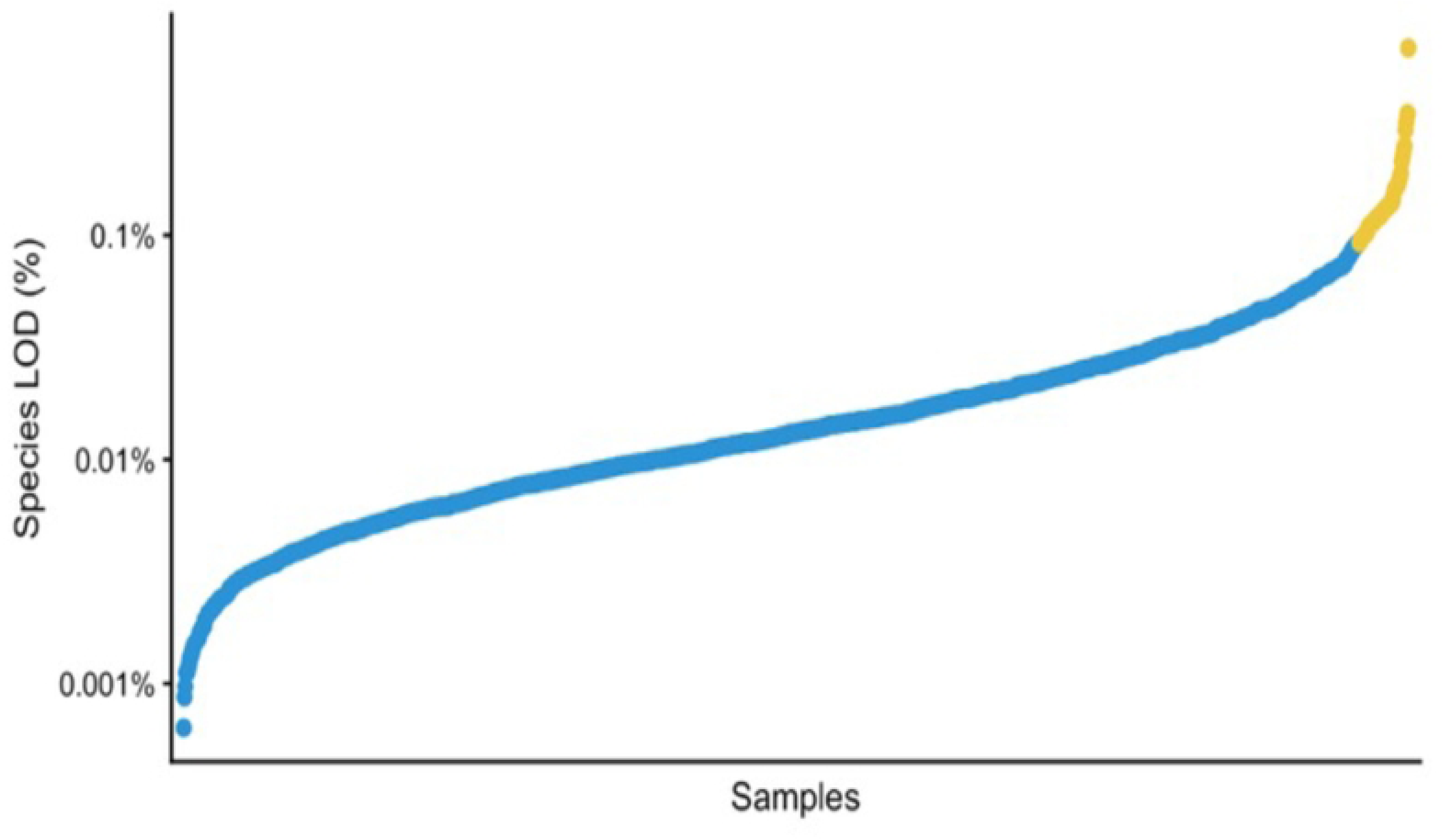
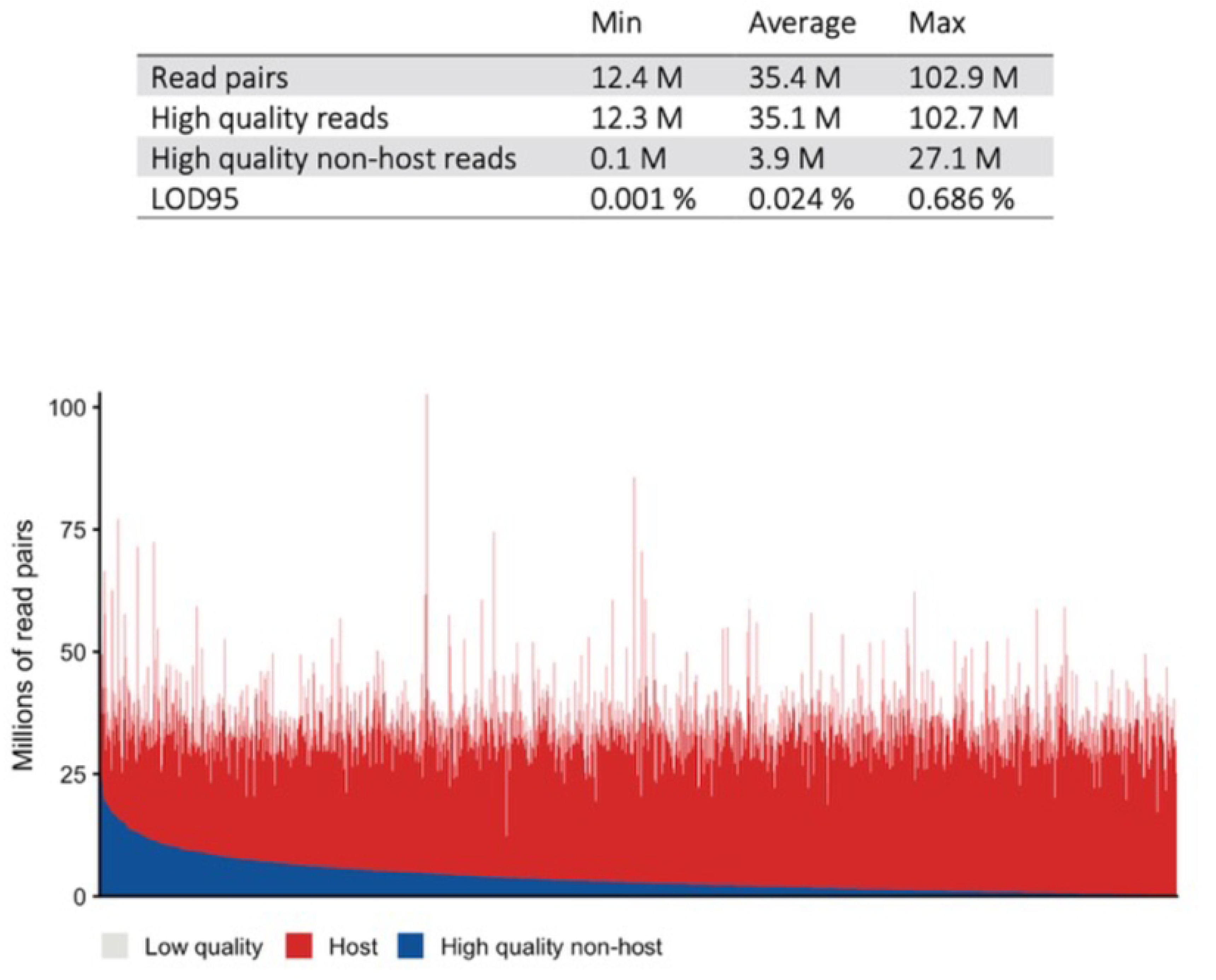
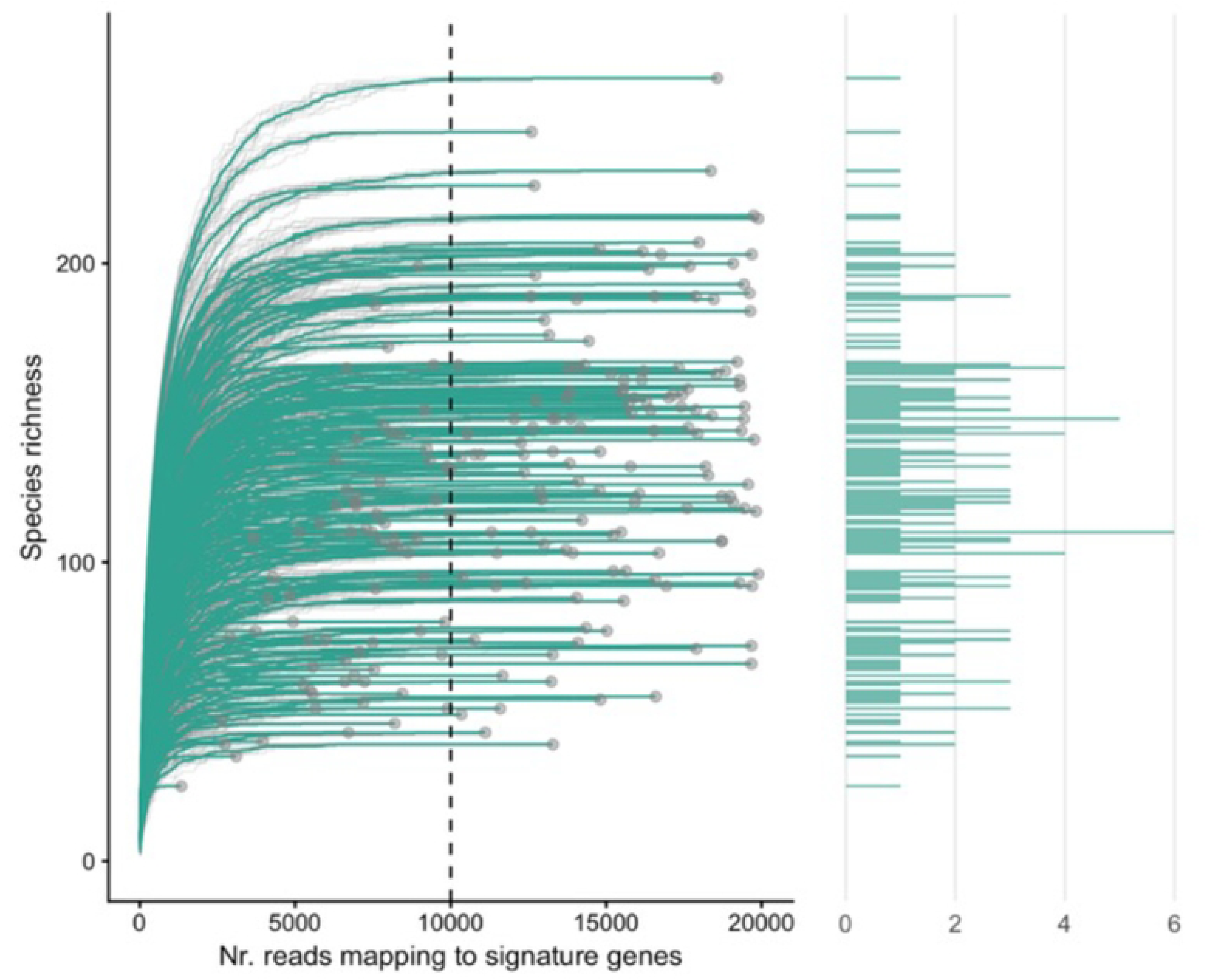
(A) Proportion of read pairs. Bar plots summarizing read quality and read mapping. Categories of different read types are given in percentages. Samples are ordered by decreasing number of mapped reads. (B) Limit of detection (LOD) of species in each sample. Dots indicate the relative abundance at which there is a 95% chance of detecting an average species. Samples are ordered by increasing LOD. Yellow dots indicate samples with a LOD above the threshold of 0.1%. (C) Bar plots summarizing read quality and read mapping for all samples. M = millions of reads. Samples are ordered by decreasing the number of mapped reads. (D) The left panel shows collector’s curves of all samples with less than 20,000 number of reads mapping to signature genes. Each curve represents one sample. The curve is drawn by randomly sampling an increasing amount of reads and estimating the richness (number of detected species). Each curve was resampled 10 times (thin grey curve) and the average is drawn in green. The dotted line represents the threshold of 10,000 reads mapping to signature genes. The right panel shows a histogram of the richness of all samples included in the left panel.

**Suppl. 3.**
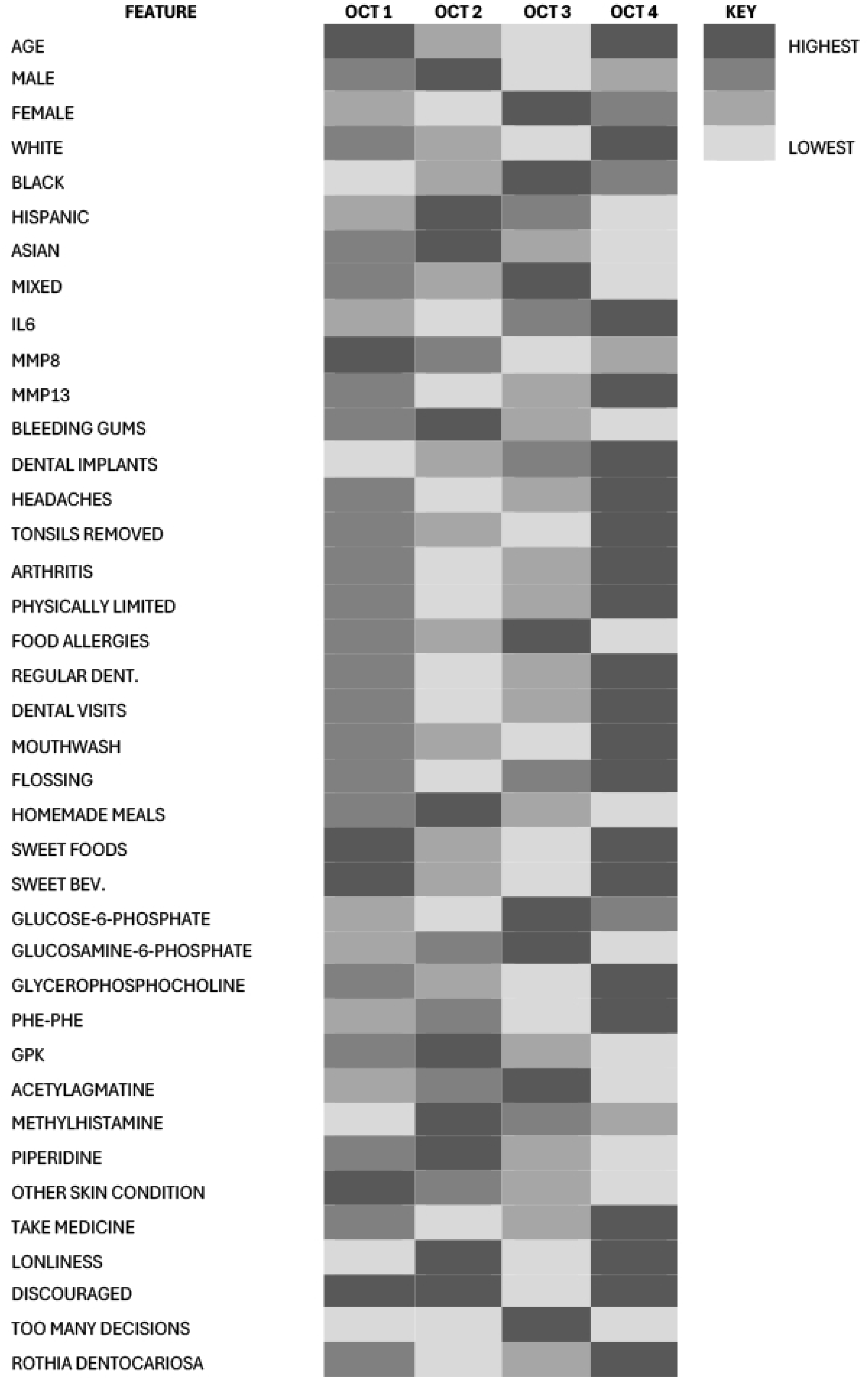
Heat Map Summary of OCTs by Feature. Heat map showing the relationship of each feature within each OCT and across the OCTs.

